# Quantitative pathology and APOE genotype reveal dementia risk and progression in Lewy body disease

**DOI:** 10.1101/2025.10.20.25338378

**Authors:** Hemanth R Nelvagal, Nancy Chiraki, Toby Curless, Patrick W Cullinane, Alice Rockliffe, Samarth Pimparkar, Haruka Kawamura, Sophie Ollerenshaw, Isha Elahi, Sebastian Brandner, Lesley Wu, Raquel Real, Mina Ryten, John Hardy, Eduardo De Pablo Fernandez, Thomas T Warner, Huw R Morris, Yau Mun Lim, Zane Jaunmuktane

**Affiliations:** Department of Clinical and Movement Neurosciences, UCL Queen Square Institute of Neurology, University College London, London, WC1N 3BG, United Kingdom; Queen Square Brain Bank for Neurological Disorders, UCL Queen Square Institute of Neurology, London, WC1N 1PJ, United Kingdom; Aligning Science Across Parkinson’s (ASAP) Collaborative Research Network, Chevy Chase, MD 20815, USA; Department of Neuromuscular Diseases, UCL Queen Square Institute of Neurology, University College London, London, WC1N 3BG, United Kingdom; Reta Lila Weston Institute of Neurological Studies, UCL Queen Square Institute of Neurology, London, WC1N 3BG, United Kingdom; Queen Square Movement Disorders Centre, UCL Queen Square Institute of Neurology, London, WC1N 3BG, United Kingdom; Department of Neurodegenerative Disease, UCL Queen Square Institute of Neurology, London, WC1N 3BG, United Kingdom; Division of Neuropathology, National Hospital for Neurology and Neurosurgery, University College London NHS Foundation Trust, London, NW1 2PB, United Kingdom; UK Dementia Research Institute at The University of Cambridge, Cambridge, CB2 0AH, United Kingdom; Centre for Preventive Neurology, Wolfson Institute of Population Health, Queen Mary University London, London, E1 4NS, United Kingdom

**Keywords:** Lewy body diseases, Parkinson’s disease, APOE genotype, Orthostatic hypotension with ischaemic pathology, Alzheimer’s co-pathology, SuStaIn

## Abstract

Dementia in Lewy body diseases (LBD) is common and arises through heterogeneous and incompletely understood pathways. Evidence suggests contributions from genetic factors, including APOE ε4 genotype, co-pathology including concomitant Alzheimer’s disease pathology and hypoperfusion related to orthostatic hypotension. However, the relative impact of these factors remains unclear. To address this, we analysed 399 post-mortem brains from LBD cases comprising Parkinson’s disease, Parkinson’s disease dementia and dementia with Lewy bodies, and controls, integrating APOE genotype, clinical data and assessment of ischaemic pathology alongside large-scale digital pathology quantification. We established an image analysis pipeline utilising machine learning to enable automated, standardised measurement of α-synuclein, amyloid-β, and phosphorylated tau burden across multiple brain regions. Quantitative pathology strongly correlated with semi-quantitative ratings and outperformed conventional staging in predicting dementia. Across multiple analytical approaches, APOE ε3 and ε4 carriers showed distinct dementia risk profiles. APOE ε3 carriers developed dementia at lower quantitative α-synuclein and amyloid-β thresholds than ε4 carriers, although overall dementia risk was dominated by ε4 genotype, consistent with ε4 both promoting greater pathology accumulation and modifying the threshold for dementia onset. Orthostatic hypotension and ischaemic pathology increased dementia risk only in ε3 carriers with low Lewy and Alzheimer’s proteinopathy burden, while male sex further modulated dementia risk for this subgroup. A data-driven progression model (SuStaIn) identified four trajectories of Lewy pathology: two corresponding to recognised patterns, one brainstem-first and the other with early amygdala involvement, and two representing novel cortical-onset patterns, one with early cingulate cortex involvement and the other starting in neocortex before limbic and brainstem involvement. Co-pathology progression modelling identified subtypes with early predominance of amyloid-β, phosphorylated tau, or α-synuclein, and showed that Lewy subtypes follow two propagation trajectories in opposite directions. Together, these findings demonstrate that integrating quantitative pathology with genotype and clinical data uncovers distinct yet overlapping pathways to dementia in LBD, refining disease progression models and providing a basis for genotype- and pathology-informed patient stratification in therapeutic trials.

## Introduction

Primary Lewy body diseases (LBD), encompass Parkinson’s disease (PD), Parkinson’s disease dementia (PDD), and dementia with Lewy bodies (DLB). Dementia is common in LBD and arises through poorly understood, heterogenous pathways. LBD represent a significant clinical and societal burden, with no disease-modifying therapies available. In PD, dementia may emerge late in the disease course, or cognitive symptoms may appear early, before or soon after motor features.^1,2^ The one-year rule classifying patients as having DLB if dementia occurs before or within a year of motor symptom onset, or PDD if dementia develops more than a year after motor onset, is clinically useful, but remains arbitrary and without a proven biological basis. Cryo-electron microscopy has shown α-synuclein filaments in PD, PDD, and DLB share identical single-protofilament structure, distinct from multiple system atrophy (MSA), highlighting both the molecular similarity within the LBD spectrum and its divergence from MSA.^3^

Several genetic and pathological factors have been implicated in LBD dementia risk. High Lewy pathology stages, increased cortical Lewy burden, and advancing Alzheimer’s disease (AD) co-pathology are associated with higher dementia risk,^4,5^ yet neuropathology cannot reliably distinguish PD, PDD and DLB in individual cases.

The APOE ε4 allele, the major genetic risk factor for AD, is also the strongest established genetic risk factor for DLB,^6^ although its effect is greatest in DLB cases with substantial AD co-pathology.^7^ APOE ε4 is not associated with risk or age of onset of PD, but is linked with higher cortical Lewy pathology^8^ and increased risk of progression to dementia in PD.^9,10^ Lewy pathology is frequently observed in AD, indicating overlapping pathophysiological processes.^11^ Experimental evidence suggests that amyloid-β (Aβ) and AD pathology can exacerbate α-synuclein aggregation,^12–14^ supporting the view that these pathologies are biologically intertwined,^15^ although the mechanisms for interaction remain unclear.

Beyond misfolded proteinopathies, orthostatic hypotension (OH), a common non-motor feature of PD, is associated with dementia,^16–18^ but neuropathological confirmation of this relationship is lacking.

Overall, the extent to which genetic, proteinopathy-related factors, OH and ischaemic pathology account for dementia risk in LBD is unclear, and additional factors are likely. Defining these processes is essential to improve prognostication, biomarker interpretation, and patient stratification for targeted therapies. To address this, we used a large post-mortem cohort of LBD and controls to generate automated quantitative measures of misfolded proteins across multiple brain regions. By integrating these with clinical, vascular and genetic data, we aimed to define how pathology patterns contribute to dementia in LBD.

## Materials and methods

### Study design and participants

This retrospective post-mortem study included neuropathologically confirmed LBD patients with clinical parkinsonism and controls without neurological disease. Cases were selected from the UCL Queen Square Brain Bank for Neurological Disorders (QSBB) archive between 1991-2022. Consecutive patients were included unless medical records were incomplete or formalin-fixed paraffin embedded (FFPE) blocks unavailable. QSBB protocols and this study have been approved by the NHS Health Research Authority Ethics Committee London-Central (REC reference 23/LO/0044). Written informed consent for the use of biological materials and clinical data in medical research was obtained for all cases from the donors or their next of kin.

### Clinical assessment

Medical records were systematically reviewed to extract demographic and clinical information, including age at death, disease duration, sex, and dementia status. Dementia was defined as the presence of objective cognitive impairment documented by a clinician or neuropsychological test severe enough to significantly affect tasks of daily living not attributable to motor impairment.^1^ The temporal relationship between motor and cognitive symptom onset was documented and applied to classify cases as PDD or DLB based on whether dementia developed more or less than one year after motor symptom onset.^2^ OH and clinical diagnosis of infarct or haemorrhage were also recorded. OH was defined as: i) documented drop in systolic / diastolic blood pressure ≥ 20 / 10 mmHg on standing,^19^ or ii) presence of postural symptoms suggestive of OH according to the impression of the treating clinician, persistent for > 6 months after therapeutic measures to address secondary (non-neurogenic) causes of OH had been implemented.^17^

### Genotyping

DNA was extracted from cerebellar or frontal lobe tissue and genotyped using the Illumina NeuroBooster array,^20^ which provides dense coverage of neurodegenerative disease–related variants. Genotype data were filtered using PLINK v2. Individuals and single nucleotide polymorphisms (SNPs) with >5% missing data were excluded using the –mind 0.05 and –geno 0.05 filters. Genotype data were imputed against the TOPMed r2 panel with Eagle v2.4 phasing on the TOPMed Imputation Server using Minimac4. APOE genotypes were inferred from imputed genotypes at the rs7412 and rs429358 SNPs, which define the ε2, ε3, and ε4 alleles. APOE ε2/ε3 and ε3/ε3 genotypes were classified as ε3, whereas ε2/ε4, ε3/ε4, and ε4/ε4 genotypes were classified as ε4. Cases carrying known pathogenic mutations in *SNCA*, *PRKN,* or *PINK1,* identified through directly genotyped or high-quality imputed variants included in the array and annotated as pathogenic or risk-associated alleles,^21^ were excluded from the study. Variants in other PD-associated risk genes, such as *GBA* and *LRRK2*, and genes linked to AD or other neurodegenerative conditions were not assessed.

### Neuropathological assessment

Archival histological slides were used where available, and new sections were prepared from archival FFPE blocks if Haematoxylin & Eosin (H&E) stained slides and slides, immunostained for α-synuclein, Aβ and phosphorylated tau (pTau), were unavailable or did not meet analytical standards. Immunostaining protocols and analysed brain regions are described in the Supplementary material. For each case, the extent of pathology was assessed according to standard criteria for Lewy (Braak stage),^22^ Aβ (Thal phase),^23^ and pTau pathology (Braak & Braak stage).^24^ Alzheimer’s disease neuropathological change (ADNC) was determined according to National Institute on Aging-Alzheimer’s Association (NIA-AA) guidelines,^25,26^ and TDP-43 pathology was evaluated using limbic-predominant age-related TDP-43 encephalopathy (LATE) staging.^27^ Pathology records were reviewed for haemorrhagic pathology and for ischaemic pathology, defined as macroscopic or microscopic infarcts, or acute ischaemic neuronal eosinophilia, in vascular watershed and other regions, recorded as present or absent.

H&E and immunohistochemistry-stained whole slides were digitised on a Hamamatsu NanoZoomer S360 (26×76 mm slides; 40× objective; JPEG compression quality 70; pixel size 0.23 µm/pixel) or Hamamatsu NanoZoomer S60 (double 52×76 mm slides; 40× objective; JPEG compression quality 65; pixel size 0.22 µm/pixel) slide scanners (Fig. 1).

**Figure 1:**
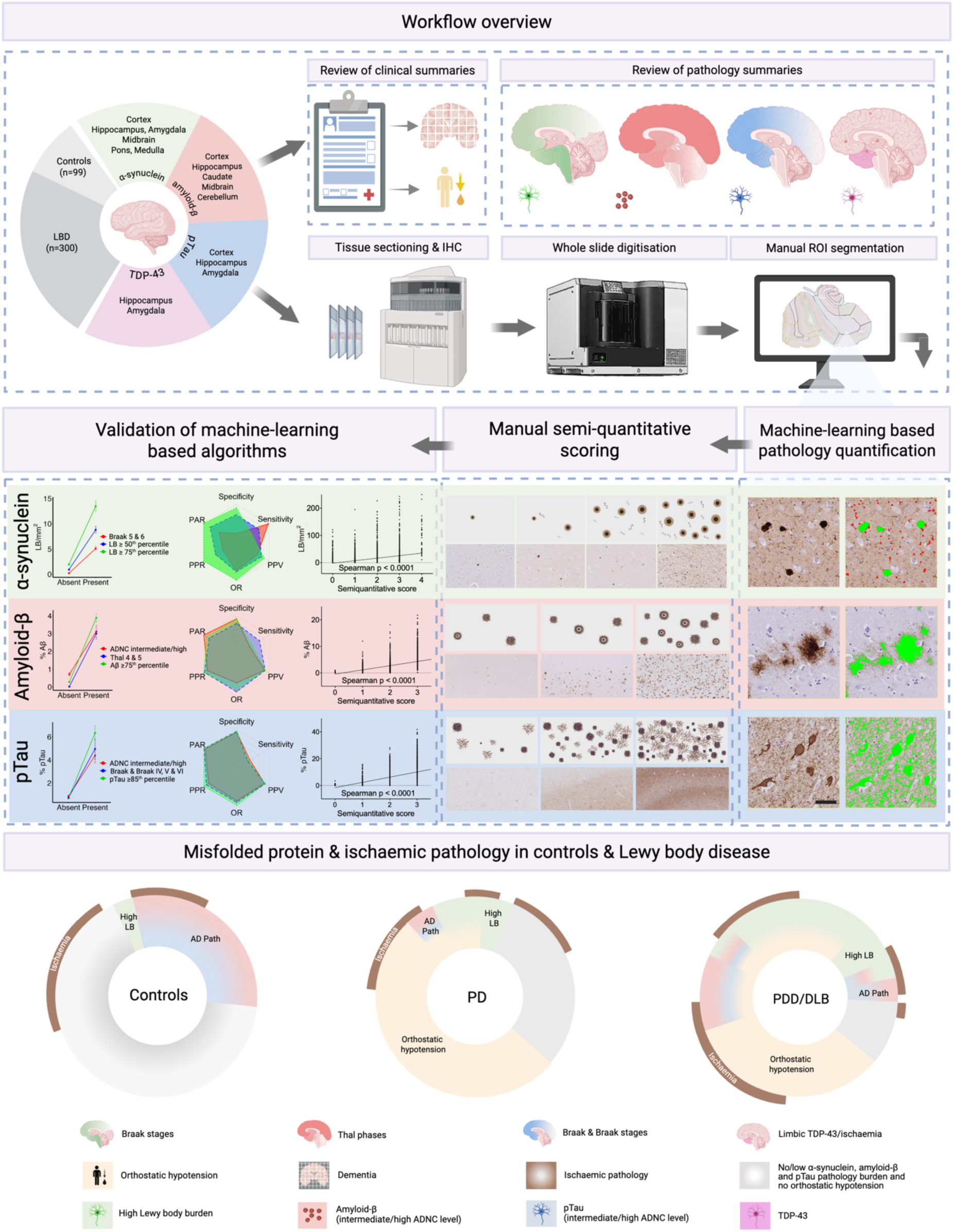
Cohort design, digital pathology quantification, method validation, and clinical correlates. **Top panel:** Schematic overview of the analytical workflow. The study included 399 cases (99 controls and 300 Lewy body disease (LBD)). Clinical summaries were reviewed for demographic information and to determine the presence of dementia, orthostatic hypotension and clinical history of stroke or haemorrhage. Pathology reports were reviewed for misfolded protein pathology, traditional pathology severity scores, and the presence of ischaemic or haemorrhagic pathology. Following digitisation of immunohistochemically (IHC) stained slides for α-synuclein, amyloid-β (Aβ), phosphorylated tau (pTau), and TDP-43, regions of interest (ROIs) were manually delineated across defined cortical, limbic and brainstem areas. **Middle panel (from right to left):** Far right, machine-learning algorithms were developed to quantify α-synuclein burden (LB/mm²), and Aβ and pTau burden (% area) across each ROI. Representative 40x images of each misfolded proteinopathy (left) with corresponding detected signal in green (right); excluded signal shown in red. Bottom panel: scale bar, 100 µm. Schematic illustration of manual semi-quantitative ratings assigned to each ROI using a 5-tier scale (0 = none, 1 = rare, 2 = occasional, 3 = moderately frequent, 4 = frequent) for α-synuclein, and a 4-tier scale (0 = none, 1 = occasional, 2 = moderately frequent, 3 = frequent) for Aβ and pTau. Spearman correlation analyses showed strong associations between machine-learning-derived quantitative pathology burden and manual semi-quantitative scores for α-synuclein, Aβ, and pTau across matched ROIs (Spearman ρ: α-synuclein = 0.72, Aβ = 0.80, pTau = 0.76; Spearman p < 0.0001 for all three pathologies). Radar plots compare performance of pathology classification methods for dementia prediction. Metrics include specificity, sensitivity, positive predictive value (PPV), odds ratio (OR), pathology presence ratio (PPR), and pathology absence ratio (PAR) (see Supplementary Table 2). Quantitative thresholds (50^th^ and 75^th^ or 85^th^ percentiles) and traditional staging criteria (Braak stage, Thal phase, Braak & Braak stage, intermediate-to-high Alzheimer’s disease neuropathological change (ADNC) levels as defined by NIA-AA criteria) are compared for each proteinopathy. Line graphs display the mean quantitative burden for each pathology, stratified by classification group and presence of dementia. Colour-coded lines and radar plots correspond to distinct pathology thresholds or staging criteria, as indicated in the key for each proteinopathy. Average pathology burden for Spearman correlation analyses, radar plots, and line graphs was calculated across the following ROIs: cortices of superior and middle frontal gyri; hippocampal subfields CA1–CA4; subiculum; parasubiculum; entorhinal and transentorhinal cortices; and cortex of the fusiform gyrus. Detailed statistical outputs for Spearman correlations, radar plot performance metrics, and quantitative burden comparisons shown in Figure 1 are available in corresponding supplementary Excel statistics data. **Bottom panel:** Circular ring plots illustrate the frequency and co-occurrence of major neuropathological and clinical features across three groups: controls, Parkinson’s disease (PD) without dementia, and combined PD with dementia (PDD) and Dementia with Lewy bodies (DLB). Each segment represents the proportion of cases with a given feature, including high (≥75th percentile) Lewy body (LB) pathology in the cortices of the superior frontal and middle frontal gyri and medial temporal regions (dentate gyrus, CA4, CA3, CA2, CA1, subiculum, para-subiculum, entorhinal cortex, transentorhinal cortex, cortex of the fusiform gyrus), Alzheimer’s disease (AD) co-pathology (intermediate-to-high, as defined by ADNC), orthostatic hypotension, ischaemic pathology, and presence of dementia (in relevant groups). Features may co-occur within individual cases and are displayed as overlapping segments. The circular ring plots allow visual comparison of the burden and overlap of neuropathological and clinical findings across the three groups. Each feature is colour-coded and symbolised as per the key below the plots.

### Quantitative digital neuropathology

For quantitative image analysis, regions of interest (ROIs) (Supplementary Table 1) were manually annotated on NZConnect (web-based) or NDP.view2 (offline) (Hamamatsu), digital whole slide image (WSI) viewing platform. In each brain region, a small circle annotation, named “!BACKGROUND” with 10.5 µm diameter was positioned where background staining was visually most intense. The annotations were downloaded as NDPA files using a custom Python script and imported into QuPath v0.5.1^28^ using a custom Groovy script for image analysis. Briefly, the ROIs were analysed by first performing colour deconvolution and background subtraction followed by applying automatic thresholding for Aβ and pTau or StarDist^29^ for Lewy bodies resulting in %Aβ, %pTau or LB/mm^2^ readouts per ROI. Detailed image analysis protocols are described in the Supplementary material.

### Semi-quantitative assessment of misfolded protein pathology

To validate quantitative pathology measures, α-synuclein, Aβ, and pTau burden were assessed semi-quantitatively by one author (NC), blinded to clinical and pathological diagnosis. Scoring was performed at 40× magnification on digitised WSIs across all ROIs on 21-inch monitor with 2560 × 1440 pixels resolution, using a 5-tier scale for α-synuclein (0 = none, 1 = rare, 2 = occasional, 3 = moderately frequent, 4 = frequent), and a 4-tier scale for Aβ and pTau (0 = none, 1 = occasional, 2 = moderately frequent, 3 = frequent), shown in Fig. 1.

### Pathology burden classification and diagnostic performance metrics

Quantitative burden of α-synuclein, Aβ and pTau in the frontal cortex and hippocampal regions was averaged and binarised into “high” and “low” groups, with thresholds set at the 50^th^ (median), 65^th^, 75^th^, 85^th^ and 90^th^ percentiles across all cases to assess association with dementia. In parallel, established neuropathological criteria were used for binary classification: cortical Lewy body presence corresponding to Braak stages 5-6, Aβ corresponding to Thal phases 4-5, pTau corresponding to Braak & Braak stages IV-VI and intermediate-to-high ADNC (Supplementary Data for Fig. 1).

To evaluate how well each pathology classification distinguished dementia status, we calculated sensitivity, specificity, odds ratio (OR), positive predictive value (PPV), pathology absence ratio (PAR), and pathology presence ratio (PPR)^30,31^ (Fig. 1, Supplementary Data, Table 1 and Supplementary Table 2). For cross-group comparability, values for OR, PAR, and PPR were normalised by dividing each by the maximum observed value across all groups for that marker.^32^ These metrics were used to generate radar plots (Fig. 1, middle section).

### Logistic regression modelling of dementia risk

To evaluate the relationship between individual predictors and dementia status, each variable was examined using univariate logistic regression models fitted with binomial family generalized linear model (glm).^33^ Analyses were stratified by APOE ε3 and ε4 genotype. Continuous predictors: α-synuclein (LB/mm²), Aβ (%Aβ), and pTau (%pTau) were entered as linear terms in the model. Categorical predictors, including sex, ischaemic pathology (as defined above), disease duration (<15 vs ≥15 years), and age at death (<77 vs ≥77 years), were entered as binary factors. 15- and 77-year cut-offs were medians for each variable within the cohort. For each model, predicted dementia probabilities were visualized using the sjPlot::plot_model() function in R.^34^ For continuous predictors, marginal effects plots were generated displaying predicted probability of dementia across the observed range of variables, with 95% confidence intervals. For categorical variables, predicted probabilities per group were plotted alongside model-derived Wald test p-values. For each APOE group, we used a fitted logistic regression model to compute the predicted probability of dementia (R code):

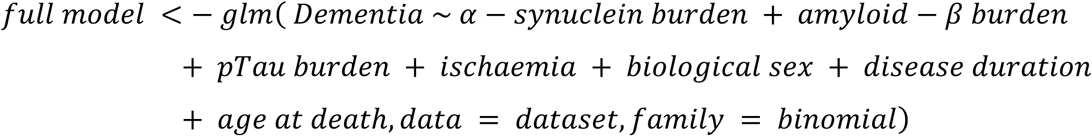

Cumulative risk heatmap and decision tree predictions (Figs 5B and 5C) are described in Supplementary material.

### Regional quantitative pathology-driven disease progression modelling

To identify distinct patterns of pathology progression, we applied the Subtype and Stage Inference (SuStaIn) algorithm^35^ to two z-score normalized datasets: (a) quantitative regional α-synuclein pathology across 11 brain regions, and (b) combined α-synuclein, Aβ, and pTau burden data from cortical and medial temporal regions, excluding amygdala. We generated synthetic control datasets, to avoid confounds from co-pathologies, demographic variability and incidental LBD in controls. Fifty synthetic controls were generated to zero-center the data, each assigned 0–5% burden across all regions and markers in both datasets. For analysis of results, only the classified LBD cases were considered. Exact SuStaIn parameters and analysed regions are described in Supplementary material.

Sankey diagrams were generated to illustrate the distribution of sex, APOE genotype, OH, ischaemic pathology, ADNC, and dementia diagnosis across subtypes (Figs 6 and 7, Supplementary Data).

### Statistical analysis

All statistical methods and statistical results are provided in the Supplementary material and supplementary Excel data files accompanying figures, tables, supplementary figures and supplementary tables respectively. A large language model (ChatGPT, OpenAI) was used to assist in generating R code for statistical analyses and data visualisation. All statistical analyses were conducted in R (version 4.4.2, RStudio version 2024.12.0.467),^36^ except for SuStaIn staging and downstream subtype analysis, which were performed in Python (Python version: 3.9.21,^37^ Jupyter Notebook version: 7.3.2).^38^ All code, statistical tests, and outputs were manually reviewed and validated by the authors to ensure methodological appropriateness and accuracy of results.

## Results

### Demographic, clinicopathological and genetic characteristics of the cohort

A total of 399 cases were included in the study, comprising 99 controls without neurological disease and 300 LBD (130 PD, 146 PDD and 24 DLB) (Fig. 1). Demographic, genetic and clinical characteristics are shown in Table 1 (and Supplementary Data). Across all disease groups, the proportion of males ranged from 55% to 83%, with no significant sex differences. APOE ε4 carrier frequency was similar in PD (24%) and PDD (26%) but was higher in DLB (55%) and lower in controls (18%) (Table 1, Supplementary Data). Age at death and disease duration were comparable between PD (76.3 ± 8.4 years; 15.9 ± 8.6 years) and PDD (77.4 ± 7.5 years; 17.7 ± 9.7 years), but both were lower in DLB (72.3 ± 6.9 years; 8.4 ± 3.2 years). OH was more frequent in PDD (72%) and DLB (88%) than PD (66%), while ischaemic pathology was increased in PDD (35%) relative to PD (27%) and DLB (25%) (Table 1, Supplementary Data).

**Table 1.** Clinical, demographic and neuropathological characteristics of study cohorts.

| Cohort | n | Male,<br>n (%) | Age at<br>onset,<br>years | Disease<br>duration,<br>years | Age at<br>death,<br>years | APOE ε4<br>carriers,<br>n (%) | Dementia,<br>n (%) | Orthostatic<br>hypotension,<br>n (%) | Ischaemic<br>pathology<br>n (%) |
| --- | --- | --- | --- | --- | --- | --- | --- | --- | --- |
| PD | 130 | 72/130<br>(55%) | 60.7 ± 11.7 | 15.9 ± 8.6 | 76.3 ± 8.4 | 26/110<br>(24%) | 0/130 (0%) | 39/59 (66%) | 35/130<br>(27%) |
| PDD | 146 | 91/146<br>(62%) | 59.8 ± 12.2 | 17.7 ± 9.7 | 77.4 ± 7.5 | 31/119<br>(26%) | 146/146<br>(100%) | 55/76 (72%) | 51/146<br>(35%) |
| DLB | 24 | 20/24 (83%) | 64.0 ± 7.3 | 8.4 ± 3.2 | 72.3 ± 6.9 | 11/20<br>(55%) | 24/24 (100%) | 7/8 (88%) | 6/24 (25%) |
| PDD+DLB | 170 | 111/170<br>(65%) | 60.4 ± 11.7 | 16.4 ± 9.6 | 76.7 ± 7.6 | 42/139<br>(30%) | 170/170<br>(100%) | 62/84 (74%) | 57/170<br>(34%) |
| Controls | 99 | 43/99 (43%) | - | - | 83.6 ± 10.8 | 10/56<br>(18%) | 0/99 (0%) | 0/14 (0%) | 33/99 (33%) |
| Cohort | n | Lewy body<br>density<br>(LB/mm<sup>2</sup>) | %Aβ | %pTau | α-synuclein<br>Braak<br>stage<sup>22</sup> | Aβ Thal<br>phase<sup>23</sup> | pTau<br>Braak &<br>Braak<br>stage<sup>24</sup> | ADNC<sup>25,26</sup> | TDP-43<br>LATE<br>stage<sup>27</sup> |
| PD | 130 | 1.147<br>[0.011–<br>21.779] | 0.054<br>[0.002–<br>5.054] | 0.278<br>[0.005–<br>7.23] | 6 [0–6] | 1 [0–5] | 2 [0–4] | 1 [0–2] | 9/79 (11%) |
| PDD | 146 | 3.922<br>[0.034–<br>34.998] | 0.453<br>[0.001–<br>7.846] | 0.65<br>[0.007–<br>18.803] | 6 [0–6] | 3 [0–5] | 2 [0–6] | 1 [0–3] | 18/107<br>(17%) |
| DLB | 24 | 7.372<br>[0.144–<br>47.453] | 1.68<br>[0.011–<br>10.623] | 1.611<br>[0.056–<br>19.313] | 6 [4–6] | 4 [0–5] | 3 [1–6] | 2 [0–3] | 3/14 (21%) |
| PDD+DLB | 170 | 4.261<br>[0.034–<br>47.453] | 0.664<br>[0.001–<br>10.623] | 0.715<br>[0.007–<br>19.313] | 6 [0–6] | 3 [0–5] | 2 [0–6] | 1 [0–3] | 21/121<br>(17%) |
| Controls | 99 | 0.026 [0–<br>6.265] | 0.446<br>[0.002–<br>11.205] | 0.632<br>[0.003–<br>11.015] | 0 [0–6] | 2 [0–5] | 2 [0–6] | 1 [0–3] | 4/66 (6%) |

Values are mean ± SD for age at onset, disease duration and age at death. Age at onset was calculated from reported symptom onset. Disease duration is symptom onset to death. Lewy body density, % immunoreactivity of amyloid-β (%Aβ), and % immunoreactivity of phosphorylated Tau (%pTau) are quantitative averages across defined cortical and medial temporal regions. Values for Lewy body density, %Aβ, and %pTau, α-synuclein Braak stage, Aβ Thal phase, pTau Braak & Braak stage and Alzheimer's disease neuropathological change (ADNC) level as per National Institute on Aging-Alzheimer's Association (NIA-AA) are median with [range]. ADNC: 0-3; Braak stage: 0-6; Aβ Thal: 0-5; pTau Braak & Braak stage: 0-VI (numbered 0-6). TDP-43 LATE staging represented as % cases, since all positive cases correspond to stage 2. "–" indicates not applicable.

APOE ε3 carriers were older at death than ε4 carriers, regardless of dementia status. Disease duration was shorter in ε4 carriers in PD and PDD (Supplementary Tables 3 and 4, Supplementary Data).

Semi-quantitative assessment showed limbic (stage 2) TDP-43 pathology in a subset of cases (Table 1), correlating with increasing age at death, but not dementia status (Supplementary Table 5, Supplementary Data). As TDP-43 pathology was not a prominent feature, automated quantification was not pursued.

### Quantitative pathology correlates with semi-quantitative scores and enhances dementia prediction

Automated digital quantification of α-synuclein, Aβ, and pTau showed strong correlations with semi-quantitative scores across frontal and hippocampal ROIs (Spearman ρ: α-synuclein = 0.72, Aβ = 0.80, pTau = 0.76; all Spearman p < 0.0001) supporting the validity of the digital pathology approach (Fig. 1, middle section, Supplementary Data).

Radar plots (Fig. 1, middle section, Supplementary Data) revealed that quantitative α-synuclein, thresholds (particularly ≥75^th^ percentile) outperformed Braak staging-based cortical Lewy pathology in dementia prediction with higher specificity, OR, and PPV. For Aβ (≥75^th^ percentile) and pTau (≥85^th^ percentile), quantitative thresholds performed similarly to Thal and Braak & Braak staging. Therefore, these thresholds were used to binarize regional quantitative pathology into ‘high’ and ‘low’ burden for all subsequent analyses (Fig. 1, Supplementary Data and Supplementary Table 2).

Line plots comparing the quantitative pathology burden showed that both quantitative score cutoffs (≥50^th^, 75^th^ or 85^th^ percentiles) and traditional staging demonstrated a progressive increase across cortical and medial temporal regions. Quantitative pathology cutoffs were consistently higher than those defined by traditional staging (Fig. 1, middle section, Supplementary Data).

In controls (Fig. 1, bottom section), a minority of individuals showed either high Lewy pathology (2/99) or at least intermediate ADNC (30/99). Most PD patients without dementia, had low Lewy and ADNC pathology (83/130). OH was frequent (39/59 with available data), and ischaemic pathology was variably present (35/130). A small subset had high Lewy burden (10/130), intermediate-to-high ADNC (6/130), or both (1/130), despite preserved cognition. Among 170 LBD dementia cases, over half showed concurrent high Lewy pathology and/or ADNC (105/170), with a large proportion of PDD/DLB cases having co-occurring OH (62/84 with available data). A minority had dementia despite low Lewy pathology and ADNC, absence of OH, and no ischaemic pathology (8/170).

### Quantitative pathology burden intensifies with advancing disease stage and APOE ε4 genotype

We examined whether quantitative regional pathology burden aligned with established staging schemes (Fig. 2, Supplementary Data) and observed that with advancing Lewy, Aβ, and pTau stage/phase, regional burdens within already affected areas also intensified - a nuance not captured by conventional staging, which documents only regional pathology presence rather than density. Lewy pathology was most pronounced in limbic and neocortical regions at Braak stage 6, Aβ burden peaked in cortical and limbic regions at Thal phase 5, and pTau in medial temporal and association cortices at Braak & Braak stage VI. Quantitative burdens of all three proteinopathies also rose with advancing ADNC: Lewy pathology most prominently in the amygdala, pTau and Aβ in limbic and neocortical regions, and Aβ additionally in the basal ganglia.

**Figure 2:**
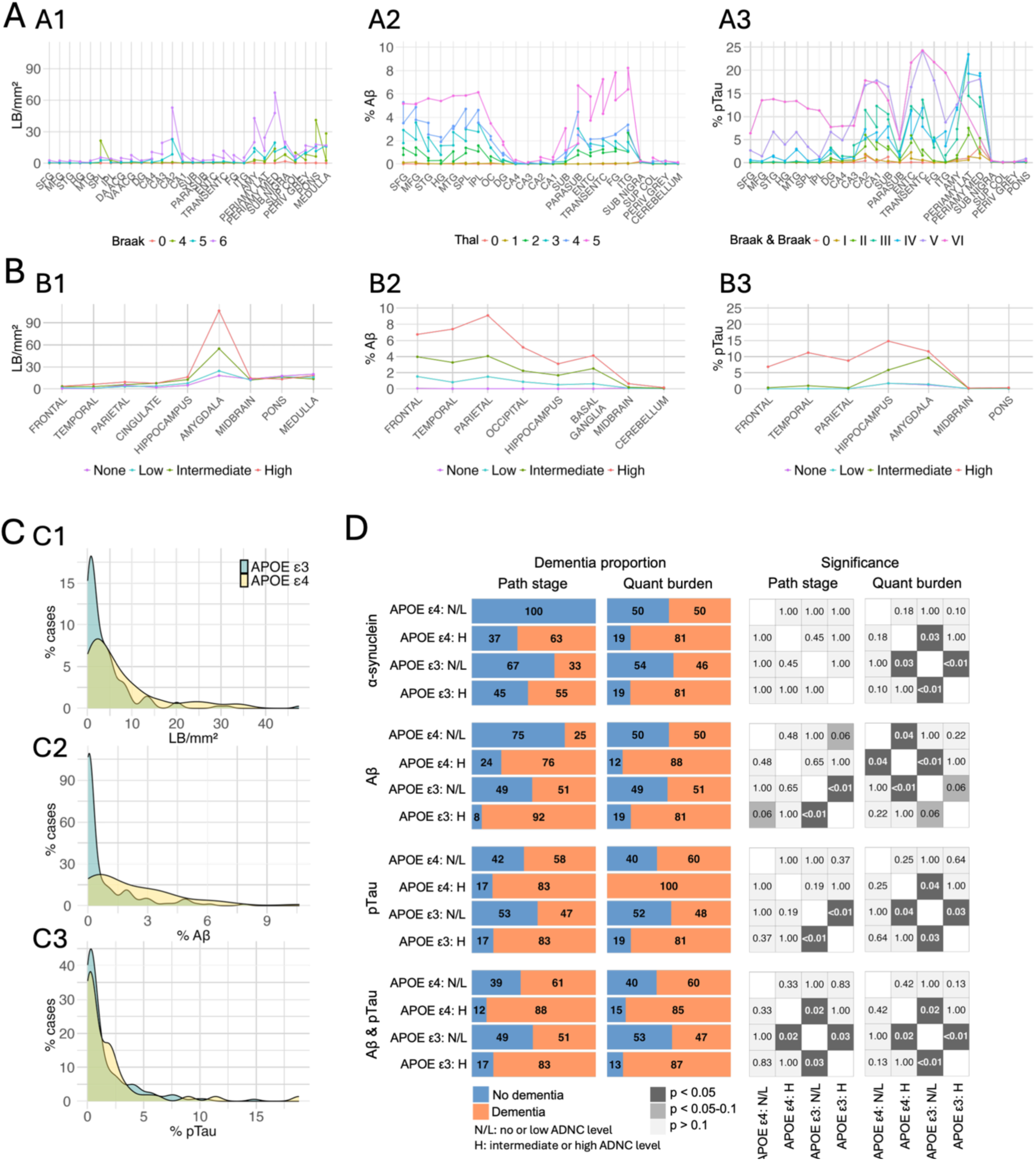
Regional quantitative pathology burden stratified by traditional staging. **(A) Quantitative pathology burden across brain regions stratified by stage or phase of the corresponding misfolded proteinopathy.** **(A1–A3)** show the quantitative pathology burden (y-axis) of Lewy bodies (**A1**, LB/mm^2^), % area of Aβ (**A2**, % Aβ), and % area of phosphorylated tau (**A3**, %pTau) across anatomical brain regions (x-axis), stratified by stage or phase of the respective proteinopathy. **A1**: Lewy body burden plotted by Braak stage (0, 4, 5, 6). **A2**: Aβ burden stratified by Thal phase (0–5). **A3**: pTau burden stratified by Braak & Braak stage (0–VI). Brain region abbreviations are detailed in Supplementary Table 1 and detailed statistical outputs are available in supplementary Excel statistics data for Figure 2A. Kruskal-Wallis tests with Dunn’s pairwise post-hoc comparisons and Benjamini-Hochberg (BH) adjustment was performed per region for each proteinopathy. **(B) Quantitative pathology burden across brain regions stratified by level of Alzheimer’s disease neuropathologic change.** **(B1–B3)** show the quantitative pathology burden (y-axis) of Lewy pathology **(B1**, LB/mm^2^), % area of Aβ (**B2**, % Aβ), and % area of phosphorylated tau (**B3**, %pTau) across anatomical brain regions (x-axis), stratified by Alzheimer’s disease neuropathological change (ADNC) level as defined by NIA-AA criteria (none, low, intermediate, high). Detailed statistical outputs are available in supplementary Excel statistics data for Figure 2B. Kruskal-Wallis tests with Dunn’s pairwise post-hoc comparisons and Benjamini-Hochberg (BH) adjustment was performed per region for each proteinopathy. Brain regions include average scores from frontal (cortices of the superior frontal gyrus and middle frontal gyrus), temporal (cortices of the superior temporal gyrus, Heschl’s gyrus and middle temporal gyrus), parietal (cortices of the superior parietal lobule and inferior parietal lobule), occipital (occipital cortex), hippocampus (dentate gyrus, CA4, CA3, CA2, CA1, subiculum, para-subiculum, entorhinal cortex, transentorhinal cortex, cortex of the fusiform gyrus), amygdala, basal ganglia (caudate nucleus and putamen), midbrain (substantia nigra, superior colliculus, periventricular grey matter), pons (pontine tegmentum), medulla (medullary tegmentum) and cerebellum. **(C) Distribution of quantitative misfolded protein pathology burdens stratified by APOE genotype.** Density plots show the percentage of cases (y-axis) across the range of measured pathology burden (x-axis) for Lewy bodies (**C1**, LB/mm^2^), % area of Aβ (**C2**, %Aβ), and % area of phosphorylated tau (**C3**, %pTau), stratified by APOE genotype. Blue represents APOE ε3 carriers; yellow represents APOE ε4 carriers. These plots visualise the frequency distribution of pathology burden across cases, highlighting variation in burden profiles by genotype. Exact two-sample Kolmogorov-Smirnov test performed for APOE ε3 vs ε4 carriers for each proteinopathy (α-synuclein p = 0.02, Aβ p = 9.7e-07, pTau p = 0.52). **(D) Comparison of pathology staging versus quantitative burden in predicting dementia frequency across APOE genotypes divided by severity of Alzheimer’s disease neuropathologic change.** **Left panels:** Bar plots show the proportion of Lewy body disease (LBD) patients with dementia (orange) versus without dementia (blue) stratified by APOE genotype (ε3 and ε4) and ADNC levels as defined by NIA-AA criteria (NL: none or low-level; H: intermediate or high-level), for each pathology type (α-synuclein, Aβ, pTau, and combined Aβ & pTau). For each, comparisons are made using either traditional pathology staging (α-synuclein: Braak stage ≥5, Aβ: Thal phase ≥4 and pTau: Braak & Braak stage ≥IV and combined Aβ/pTau: intermediate/high ADNC (Path stage)) or quantitative pathology burden (α-synuclein: ≥ 75^th^ percentile, Aβ: ≥ 75^th^ percentile and pTau: ≥ 85^th^ percentile (Quant burden)). **Right panels:** Heatmaps show the statistical significance of dementia frequency differences between APOE/ADNC groups for each classification approach, assessed using Chi-square tests or Fisher’s exact tests (when counts <5) with Bonferroni post-hoc correction. p-values are visualised by shading: p < 0.05 (black), p = 0.05–0.1 (dark grey), p > 0.1 (light grey). In **C** and **D** significant quantitative pathology burden was defined as: ≥75^th^ percentile for α-synuclein (LB/mm²) and Aβ (% area); ≥85^th^ percentile for pTau (% area) and pathology burden was averaged across the following regions: cortices of the superior and middle frontal gyri, CA1– CA4, subiculum, parasubiculum, entorhinal cortex, transentorhinal cortex, and cortex of the fusiform gyrus. Detailed statistical outputs are available in supplementary Excel statistics data for Figure 2D.

APOE ε4 carriers showed a broader distribution of α-synuclein, Aβ, and pTau pathology burden and more cases with high burden than ε3 carriers. In frontal and limbic regions, density curves for ε4 carriers shifted rightward, indicating greater accumulation; this shift was significant for Lewy pathology and Aβ but not for pTau (Fig. 2C, Supplementary Data).

Quantitative measures of Lewy, Aβ, and pTau burden in frontal and medial temporal regions identified stronger group differences in dementia risk than traditional staging. Dementia proportion was higher in APOE ε4 carriers and those with intermediate-to-high ADNC (Fig. 2D, Supplementary Data). Quantitative Lewy burden discriminated dementia risk more effectively than Braak cortical stages 5–6. Similarly, quantitative Aβ and pTau measures showed greater group differences than Thal phases 4-5 and Braak stages V-VI, with comparable discrimination when Aβ and pTau measures were combined and contrasted to intermediate-high ADNC.

### APOE genotype modifies regional pathology burden in relation to dementia status

Next, we compared regional pathology burdens, stratified by APOE genotype and dementia status (Fig. 3, Supplementary Data). In PD without dementia, Lewy pathology did not differ by genotype except for higher burden in superior temporal gyrus in ε4 carriers (p < 0.05). In PDD/DLB, ε4 carriers showed greater Lewy pathology across cortical and medial temporal regions, but not in brainstem (Supplementary Fig. 1A, Supplementary Data). Aβ showed broader genotype effects: ε4 carriers had higher cortical and subicular burden even without dementia, with more widespread differences in PDD/DLB. APOE ε4 carriers without dementia exhibited Aβ levels comparable to ε3 carriers with dementia (Supplementary Fig. 1B, Supplementary Data). pTau burden did not differ significantly by genotype, though it was elevated in cortical/limbic regions for ε3 carriers and in hippocampal areas for ε4 carriers with PDD/DLB (Supplementary Fig. 1C, Supplementary Data).

**Figure 3:**
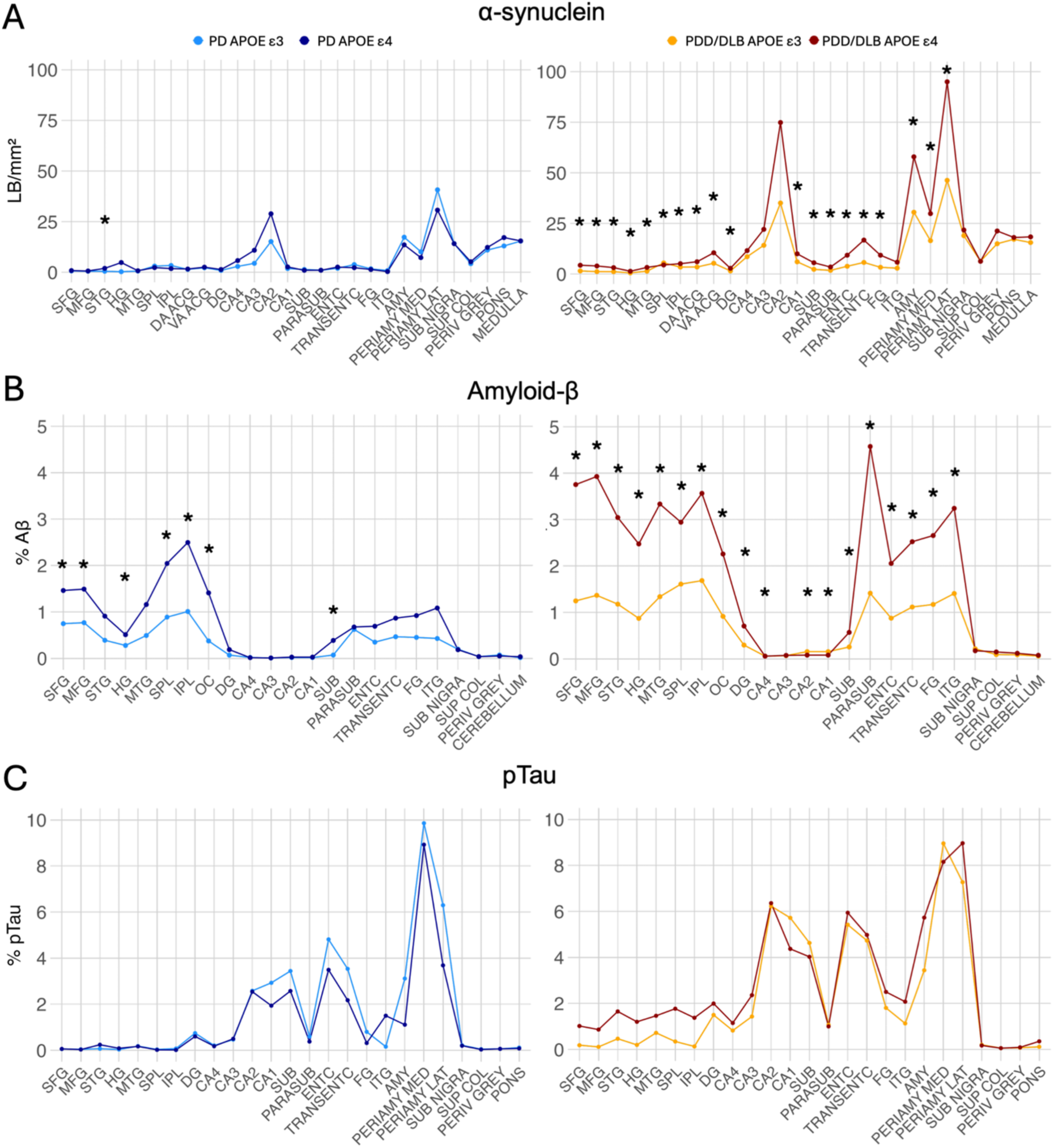
Influence of APOE genotype on regional α-synuclein, Aβ, and pTau pathology in Lewy body disease with and without dementia. **(A)** Lewy body (LB/mm^2^), **(B)** % area of Aβ (%Aβ) and **(C**) % area of phosphorylated tau (%pTau) quantitative pathology burden across multiple brain regions in Parkinson’s disease (PD) and dementia groups: Parkinson’s disease with dementia (PDD) and dementia with Lewy bodies (DLB), comparing APOE ε3 (light blue in PD, yellow in PDD/DLB) and APOE ε4 (dark blue in PD, brown in PDD/DLB) carriers. Lines indicate group mean pathology burden across regions; * indicates statistically significant differences between genotype groups in each region (p < 0.05; Mann-Whitney test with BH correction per region for each proteinopathy). Brain region abbreviations are detailed in Supplementary Table 1 and detailed statistical outputs are available in supplementary Excel statistics data for Figure 3.

### Orthostatic hypotension and ischaemic pathology elevate dementia risk at low misfolded protein burden

As OH is common in LBD and reduced cerebral perfusion may contribute to ischaemia, we examined their overlap. Among dementia cases, 69% had both OH and ischaemic pathology versus 30% without dementia (Fig. 4A). The proportions with OH alone, ischaemic pathology alone, or neither were similar across groups. When stratified by combined misfolded protein burden, ischaemic pathology was linked to a non-significant trend towards higher dementia risk in low-burden cases, whereas in high-burden cases dementia risk was elevated irrespective of ischaemic pathology (Fig. 4B, Supplementary Data).

**Figure 4:**
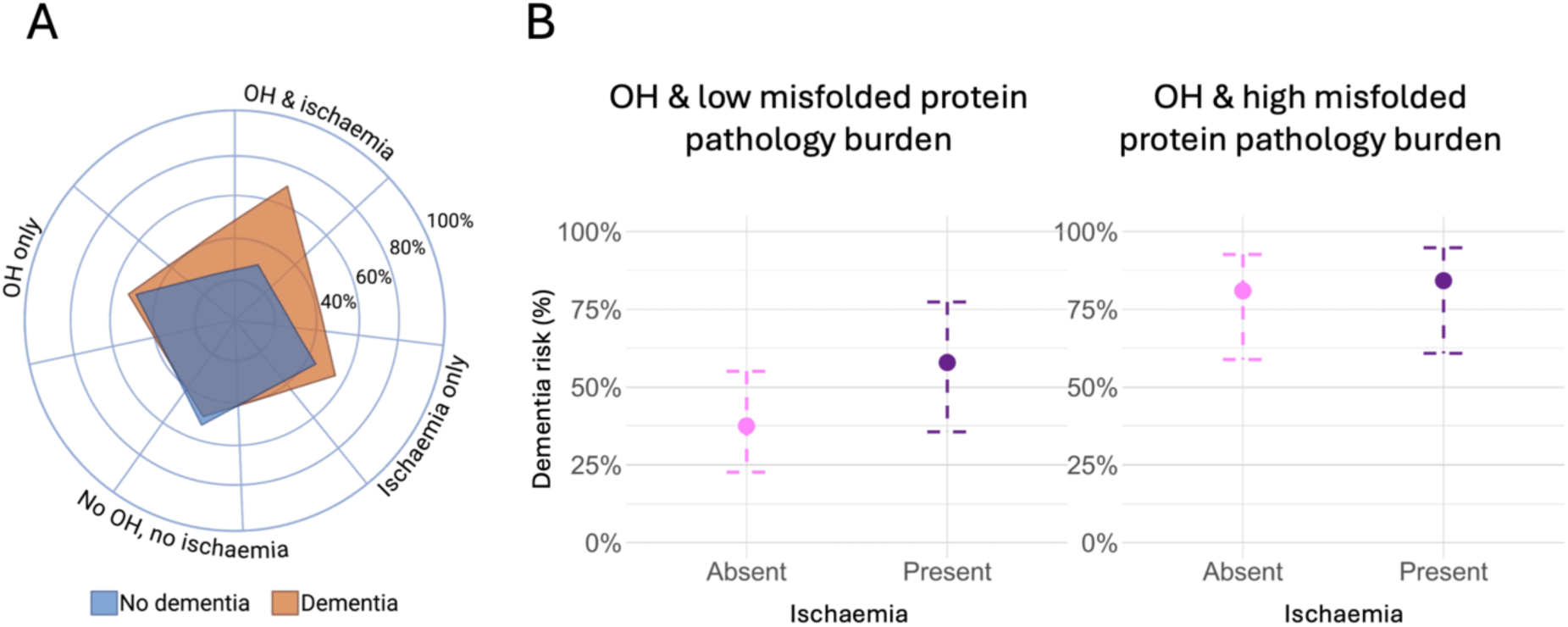
Contribution of orthostatic hypotension and ischaemic pathology to dementia risk in Lewy body disease. **(A)** Radar plot demonstrating the proportion of Lewy body disease patients with (orange) and without (blue) dementia, stratified by presence of orthostatic hypotension (OH), histological evidence of ischaemic brain pathology, both OH and ischaemic pathology, and neither. **(B)** Dot-and-whisker plots show dementia risk in patients with OH stratified by combined misfolded protein burden (low vs high, defined by composite quantitative burden of α-synuclein, Aβ, and pTau) and presence or absence of histologically confirmed ischaemic pathology. In the group with OH and low misfolded protein burden, the presence of ischaemic pathology is associated with a trend toward increased dementia risk (OR = 2.29, 95% CI = 0.72-7.30, p = 0.161), whereas in those with high misfolded protein burden, dementia risk is uniformly high regardless of ischaemic pathology (OR = 1.25, 95% CI = 0.24, 6.50, p = 0.787); analysis performed with binary logistic regression. High vs low pathology burden was defined using the ≥75^th^ percentile for α-synuclein and Aβ, and ≥85^th^ percentile for pTau. Quantitative averages were calculated across the frontal cortex (superior and middle frontal gyri) and medial temporal regions, including hippocampal subfields (CA1–CA4, dentate gyrus, subiculum, parasubiculum), entorhinal and transentorhinal cortices and cortex of the fusiform gyrus.

### Multivariable modelling reveals distinct predictors of dementia

We applied multivariable logistic regression to assess independent associations of misfolded proteinopathies and covariates (ischaemia, sex, disease duration, age at death) with dementia risk, stratified by APOE ε3 and ε4 (Fig. 5A). Lewy pathology predicted dementia in both genotypes; Aβ only in ε4; pTau in neither. Ischaemia was associated with dementia in ε3, while sex, disease duration, and age at death were not significant (Fig. 5A, Supplementary Data). To examine how misfolded protein, vascular, clinical and demographic variables interact, we generated cumulative risk heatmaps stratified by APOE (Fig. 5B, Supplementary Data). Heatmaps showed that in APOE ε4 carriers, dementia probability was high across most contexts even at low pathology burdens. In ε3, ischaemia and male sex contributed substantially to elevated risk, whereas in ε4 carriers these modifiers had minimal impact due to already near-saturated probabilities (Supplementary Data for Fig. 5B).

**Figure 5:**
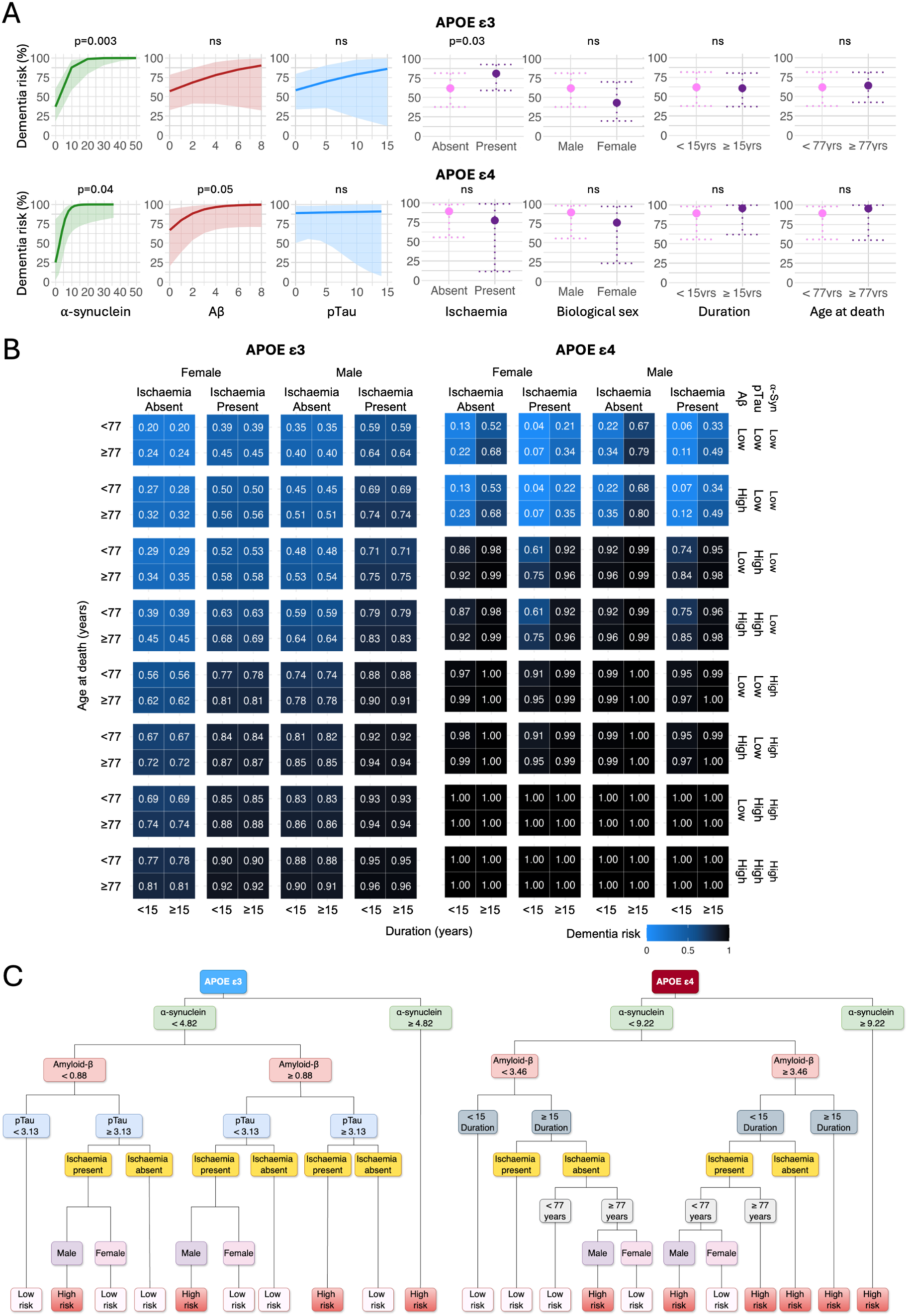
Differential predictors of dementia risk in Lewy body disease: impact of APOE genotype, misfolded protein, ischaemic pathology, demographics, and clinical features. **(A)** Predicted dementia probabilities for individual variables from multivariate logistic regression models, stratified by APOE ε3 (top, n= 126) and ε4 (bottom, n= 46) genotype. Left three plots in each row display predicted probabilities as a function of increasing α-synuclein (green) (APOE ε3: log(OR) = 0.25, 95% CI 0.10–0.43, p = 0.003, APOE ε4: log(OR) = 0.43, 95% CI 0.12–0.92, p = 0.040), Aβ (red) (APOE ε3: log(OR) = 0.24, 95% CI –0.11–0.67, p = 0.203, APOE ε4: log(OR) = 0.66, 95% CI 0.10–1.60, p = 0.05), and phosphorylated tau (pTau, blue) (APOE ε3: log(OR) = 0.10, 95% CI –0.16–0.38, p = 0.462, APOE ε4: log(OR) = 0.02, 95% CI –0.29–0.64, p = 0.928) pathology burden. Dark lines and shaded bands indicate model-estimated means and 95% confidence intervals. Right four plots show predicted dementia probability across binary covariates: ischaemic pathology (present/absent) (APOE ε3: log(OR) = 0.96, 95% CI 0.07–1.80, p = 0.035, APOE ε4: log(OR) = −0.88, 95% CI –3.70–1.90, p = 0.538), biological sex (male/female) (APOE ε3: log(OR) = 0.76, 95% CI –0.11–1.60, p = 0.088, APOE ε4: log(OR) = 0.96, 95% CI –1.10–3.10, p = 0.372), disease duration (<15 vs ≥15 years) (APOE ε3: log(OR) = −0.05, 95% CI –0.91–0.81, p = 0.909, APOE ε4: log(OR) = 0.99, 95% CI –1.00–3.00, p = 0.330), and age at death (<77 vs ≥77 years) (APOE ε3: log(OR) = 0.10, 95% CI –0.79–0.98, p = 0.833, APOE ε4: log(OR) = 0.91, 95% CI –1.20–3.10, p = 0.407). Large circles and dotted error bars indicate model-estimated group means and 95% confidence intervals. All effects were adjusted within genotype-specific multivariate model using multivariate logistic regression with linear terms, and Wald tests were applied on model coefficients. **(B)** Cumulative dementia risk heatmaps for APOE ε3 and ε4 genotypes, showing predicted probabilities of dementia (0-1 (absent-present), colour-coded from blue to black) for all combinations of clinical, demographic, and dichotomised pathology burden variables. Significant pathology burden was defined using the ≥75^th^ percentile for α-synuclein and Aβ, and ≥85^th^ percentile for pTau. Quantitative averages were calculated across the frontal cortex (superior and middle frontal gyri) and medial temporal regions, including hippocampal subfields (CA1–CA4, dentate gyrus, subiculum, parasubiculum), entorhinal and transentorhinal cortices and cortex of the fusiform gyrus. Detailed probability outputs are available in supplementary Excel statistics data for Figure 5B. **(C)** Decision trees showing hierarchical and threshold-based contributions of pathology and clinical variables to dementia risk for APOE å3 (left) and å4 (right) carriers. Tree splits use fixed, genotype-specific thresholds for the continuous pathology measures: Lewy body density (LB/mm²; α-synuclein), Aβ % area, and pTau % area. Within each APOE subgroup, ≥75^th^ percentile is used for α-synuclein and Aβ, and ≥85^th^ percentile for pTau. These fixed cut-points are applied consistently across all cross-validation folds. Additional splits are based on binary ischaemic pathology (absent/present) and demographic/clinical variables (biological sex, age at death (<77 vs ≥77 years), and disease duration (15< vs ≥15 years)). Terminal nodes represent the predicted probability of dementia. All models included α-synuclein, Aβ and pTau quantitative burden, ischaemic pathology, biological sex, age at death, and disease duration as input features. Analyses were conducted on Lewy body disease carriers with APOE ε3 (n = 181) and APOE ε4 (n = 69).

Decision-tree models were fitted separately for APOE ε3 carriers (n = 181) and ε4 carriers (n = 69) (Fig. 5C). Split thresholds for α-synuclein, Aβ, and pTau were pre-defined from within-group distributions and held constant across training folds, ensuring consistent and interpretable “high” vs “low” definitions. Cross-validated areas under curves (AUCs) were 0.879 (ε3) and 0.923 (ε4), with optimal probability thresholds of 0.562 (ε3) and 0.583 (ε4), yielding sensitivity/specificity of 0.729/0.839 (ε3) and 0.767/0.938 (ε4), respectively. Thus, both models achieved high accuracy, with ε4 showing stronger discrimination (notably specificity) and ε3 a more balanced sensitivity-specificity trade-off. α-Synuclein burden was the strongest dementia predictor in both genotypes, with thresholds of 4.82 LB/mm² (ε3) and 9.22 LB/mm² (ε4). Aβ thresholds were 0.88% area (ε3) and 3.46% area (ε4). In ε3 carriers, pTau ranked third, followed by ischaemia and sex; in ε4 carriers, disease duration ranked third, followed by ischaemia, age at death, and sex. pTau contributed only in ε3, while age at death and disease duration contributed only in ε4. Male sex increased dementia probability in both groups.

### Data-driven modelling of Lewy pathology reveals four subtypes with clinical and genetic associations

To examine how pathological variables relate to Lewy pathology progression, we applied SuStaIn using quantitative Lewy burden from brainstem, limbic, and cortical regions (Fig. 6, Supplementary Fig. 2). In 295 LBD cases, a single-subtype model inferred a canonical sequence beginning in the brainstem, spreading to the amygdala and anterior cingulate, and ultimately involving neocortex. Subtype analysis identified four distinct trajectories: (1) amygdala subtype (n = 127, 43%) with early amygdala and brainstem involvement; (2) brainstem subtype (n = 94, 31.8%) with early brainstem-restricted pathology; (3) cingulate subtype (n = 46, 15.6%) with early anterior cingulate and brainstem involvement; and (4) neocortical subtype (n = 28, 9.5%) with early frontal/temporal involvement followed by cingulate, amygdala, and brainstem at later stages.

**Figure 6:**
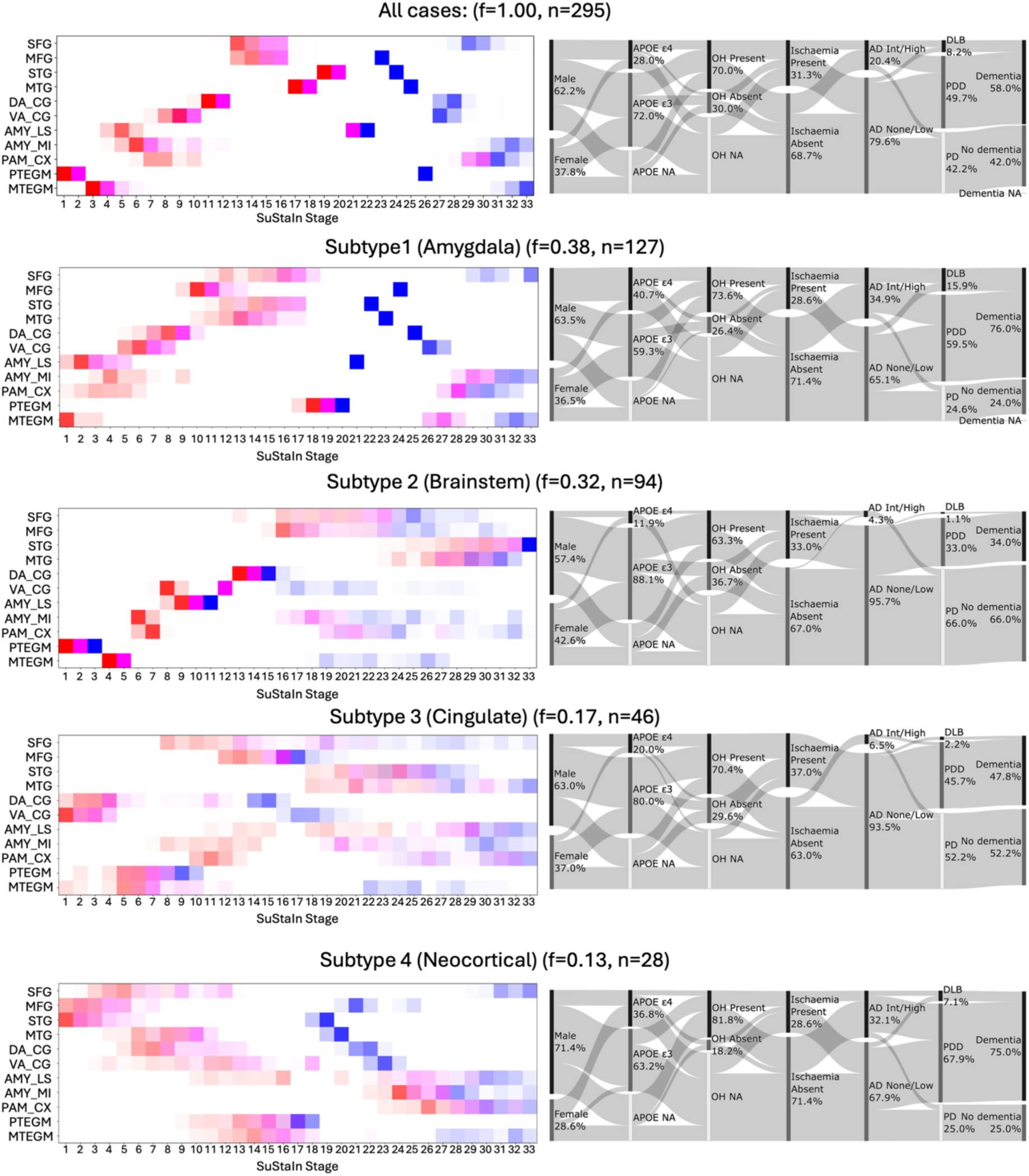
Data-driven trajectories of Lewy pathology and associated clinical features in primary Lewy body disease. The left panel shows SuStaIn-inferred positional variance diagrams with spatiotemporal trajectories of Lewy pathology progression based on the quantified Lewy body burden (LB/mm²). The horizontal axis represents predicted stages of regional involvement, while the vertical axis displays anatomical regions: cortex of the superior frontal gyrus (SFG) and middle frontal gyrus (MFG), cortex of the superior temporal gyrus (STG) and middle temporal gyrus (MTG), dorsal anterior cingulate cortex (DA_CG) and ventral anterior cingulate cortex (VA_CG), lateral aspect of the amygdala (AMY_LS) and medial aspect of the amygdala (AMY_MI), periamygdaloid cortex (PAM_CX), pontine tegmentum (PTEGM) and medulla tegmentum (MTEGM). Colours indicate the probability of a region reaching abnormality thresholds relative to controls at each disease stage: mild (red), moderate (magenta), and severe (blue). Colour intensity reflects model certainty, with deeper hues indicating higher certainty that the biomarker is abnormal at that stage, paler shades indicating greater uncertainty (higher positional variance), and white representing minimal or no probability. For each subtype, “f” represents the model-estimated, likelihood-weighted frequency across all Markov Chain Monte Carlo iterations, whereas “n” indicates the number of individuals with Lewy body disease assigned to that subtype. When modelled as a single type (n=295), SuStaIn infers a stereotyped progression of Lewy body pathology starting in the brainstem, followed by the amygdala, anterior cingulate cortex, and neocortical regions. Subtyping reveals four distinct trajectories: Subtype 1 (Amygdala) (n = 127): early involvement of the amygdala and brainstem. Subtype 2 (Brainstem) (n = 94): pathology restricted to the brainstem at early stages. Subtype 3 (Cingulate) (n = 46): early involvement of anterior cingulate cortex and brainstem. Subtype 4 (Neocortical) (n = 28): early pathology in cortical regions, with much later amygdala and brainstem involvement. The right panel displays Sankey plots showing associations between each SuStaIn-derived subtype and clinicopathological variables, including biological sex, APOE genotype (ε3, ε4), orthostatic hypotension (OH), ischaemic pathology, Alzheimer’s disease neuropathological change (ADNC) levels, as per NIA-AA criteria (AD None/Low: none or low ADNC level; AD Int/High: intermediate or high ADNC level), and dementia status. Flow width is proportional to the percentage of cases per feature combination excluding cases with unavailable data. Detailed statistical outputs for Chi-square tests are available in supplementary Excel statistics data for Figure 6.

Sankey analysis revealed distinct genetic and clinical profiles. The amygdala subtype was enriched for APOE ε4 carriers (40.7%) and had the highest proportion of AD co-pathology (34.9%). The neocortical subtype also had a higher-than-expected proportion of APOE ε4 carriers (36.8%), AD co-pathology (32.1%) and showed the highest OH frequency (81.8%). Both amygdala (76%) and neocortical (75%) subtypes showed higher dementia prevalence and had the highest proportion of cases diagnosed as DLB (amygdala subtype:15.9%, neocortical subtype: 7.1%). In contrast, the brainstem subtype was enriched for APOE ε3 carriers (88.1%), showed low AD co-pathology (4.3%), and had markedly lower dementia prevalence (34%), although dementia occurred in a proportion of cases without significant AD co-pathology (35.5% of cases with low AD co-pathology). The cingulate subtype showed intermediate characteristics across variables (Fig. 6).

Pairwise comparisons using Chi-square tests with Bonferroni correction revealed pronounced differences between amygdala and brainstem subtypes, with significant variation in APOE genotype (adj. p = 0.017), AD co-pathology burden (adj. p = <0.0001), DLB/PDD/PD diagnosis (adj. p = <0.0001), and dementia prevalence (adj. p = <0.0001). Significant differences were observed for AD co-pathology between brainstem and neocortical subtypes (adj. p = 0.0088) and amygdala and cingulate subtypes (adj. p = 0.0040); for DLB/PDD/PD diagnosis between brainstem and neocortical subtypes (adj. p = 0.0138) and between amygdala and cingulate subtypes (adj. p = 0.0321); and for dementia prevalence between brainstem and neocortical subtypes (adj. p = 0.0123). These findings indicate that amygdala and brainstem subtypes are the most distinct in their genetic, pathological, and clinical profiles (Supplementary Data for Fig. 6). While, amygdala and brainstem subtypes represent the most common (∼75%) and distinct phenotypes, the minority of other subtypes capture atypical but clinically relevant progression patterns.

Regional quantitative pathology burdens differed across SuStaIn-derived α-synuclein subtypes (Supplementary Fig. 3). The amygdala subtype showed the greatest cortical involvement and highest limbic Aβ and pTau. The brainstem subtype showed minimal cortical pathology and low Aβ/pTau. The cingulate subtype had moderate α-synuclein in parietal/cingulate regions with modest medial temporal Aβ/pTau, while the neocortical subtype was defined by high cortical Aβ, modest pTau, and limited α-synuclein outside temporal cortex. These findings highlight heterogeneity in co-pathology and progression patterns across Lewy subtypes (Fig. 6).

### SuStaIn modelling of α-synuclein, Aβ and pTau identifies distinct APOE and co-pathology trajectories

Application of SuStaIn to 300 LBD cases using quantitative cortical and medial temporal, excluding amygdala, measures of α-synuclein, Aβ, and pTau revealed Lewy pathology as the dominant driver and identified four distinct progression subtypes with differing temporal patterns (Fig. 7, Supplementary Fig. 4). Subtype 1 (“Aβ”, n = 133, 44.3%) showed early cortical Aβ, followed by cortical then medial temporal α-synuclein. Subtype 2 (“α-synuclein 1”, n = 83, 27.6%) resembled the classical pattern, with medial temporal α-synuclein preceding cortical involvement, medial temporal pTau emerging later, and cortical Aβ last. Subtype 3 (“pTau”, n = 56, 18.6%) was characterised by early medial temporal pTau, followed by cortical and then medial temporal α-synuclein. Subtype 4 (“α-synuclein 2”, n = 28, 9.3%) was also α-synuclein-driven but showed an atypical trajectory with initial parietal cortex involvement before medial temporal spread (Fig. 7, Supplementary Fig. 4).

**Figure 7:**
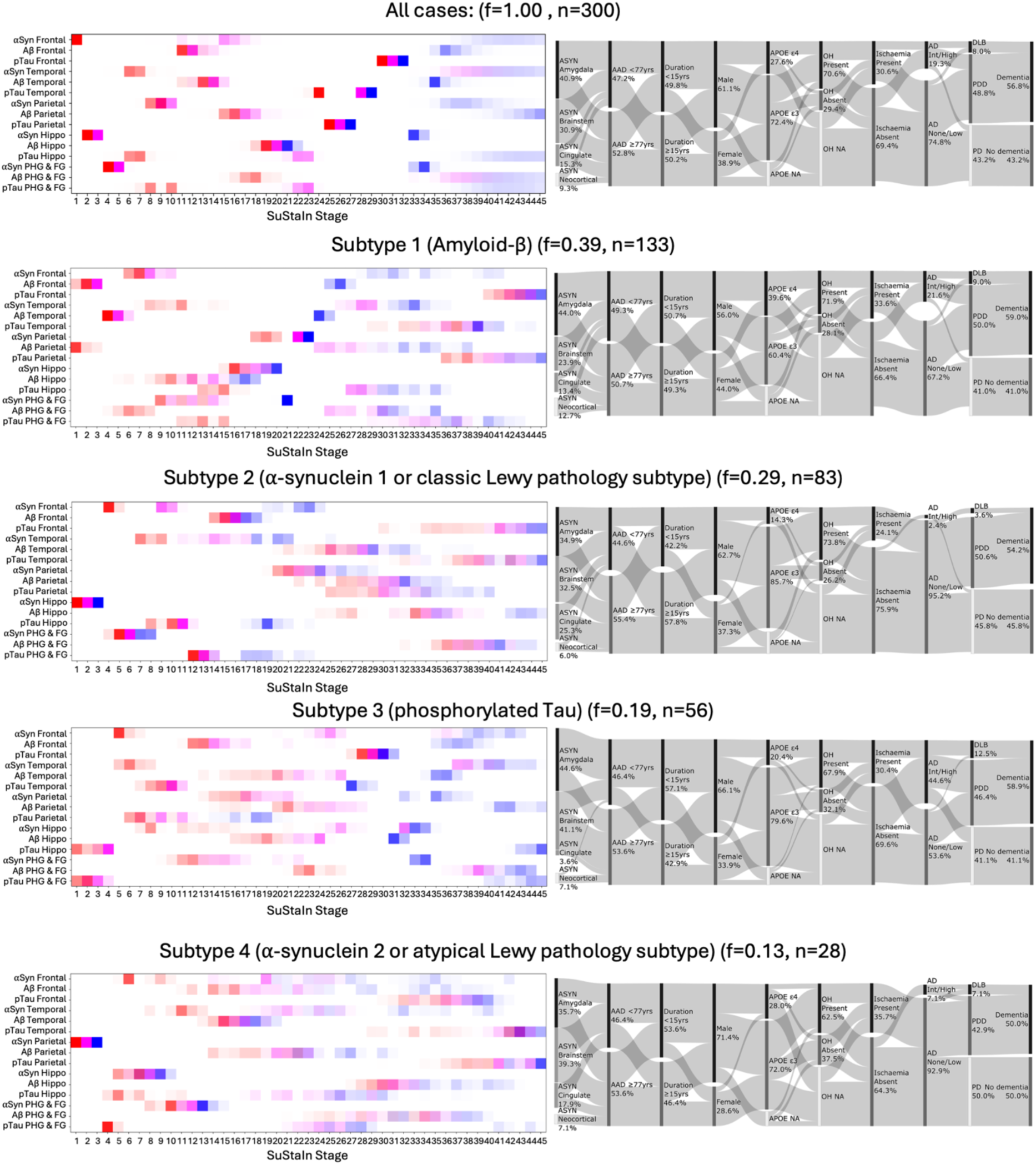
Combined data-driven modelling of α-synuclein, Aβ, and pTau pathology progression in primary Lewy body disease. The left panel shows SuStaIn-inferred spatiotemporal trajectories of disease progression based on the quantified Lewy body burden (Lewy bodies, LB/mm²), Aβ (% area), and phosphorylated tau (pTau, % area). The horizontal axis represents predicted stages of pathology regional involvement, while the vertical axis displays proteinopathies and anatomical regions: parasubiculum, entorhinal and transentorhinal cortex and cortex of the fusiform gyrus (PHG&FG); hippocampal regions, including dentate gyrus, CA4, CA3, CA2, CA1 and subiculum (Hippo); parietal cortex; temporal cortex and frontal cortex. Colours indicate the probability of a region reaching abnormality thresholds relative to controls at each disease stage: mild (red), moderate (magenta), and severe (blue). Colour intensity reflects model certainty, with deeper hues indicating higher certainty that the biomarker is abnormal at that stage, paler shades indicating greater uncertainty (higher positional variance), and white representing minimal or no probability. For each subtype, “f” represents the model-estimated, likelihood-weighted frequency across all Markov Chain Monte Carlo iterations, whereas “n” indicates the number of individuals with Lewy body disease assigned to that subtype. When modelled as a single type (n=300), SuStaIn assigns the importance to early and dominant α-synuclein pathology. Subtyping identifies four distinct progression subtypes: Subtype 1 (Amyloid-β): early cortical Aβ accumulation followed by cortical α-synuclein pathology (n = 133). Subtype 2 (α-synuclein 1 or classic Lewy pathology subtype): early medial temporal α-synuclein pathology followed by frontal and temporal involvement (n = 83). Subtype 3 (phosphorylated Tau): early medial temporal tau pathology followed by frontal and temporal α-synuclein pathology (n = 56). Subtype 4 (α-synuclein 2 or atypical Lewy pathology subtype): early parietal cortex α-synuclein pathology followed by medial temporal pTau and α-synuclein pathology (n = 28). The right panel displays Sankey plots showing associations between each SuStaIn-derived subtype and clinicopathological variables, including α-synuclein SuStaIn-derived subtypes (Figure 6), age at death (AAD) (<77 vs ≥77 years), disease duration (<15 vs ≥15 years), biological sex, APOE genotype (ε3, ε4), orthostatic hypotension (OH), ischaemic pathology, Alzheimer’s disease neuropathological change (ADNC) levels, as per NIA-AA criteria (AD None/Low: none or low ADNC level; Int/High: intermediate or high ADNC level) and dementia status. Flow width is proportional to the percentage of cases per feature combination excluding cases with unavailable data. Detailed statistical outputs for Chi-square tests are available in supplementary Excel statistics data for Figure 7.

Sankey visualisation showed that the “Aβ” subtype had the highest proportion of APOE ε4 carriers (39.6%) and elevated intermediate-to-high ADNC (21.6%). The “α-synuclein 1” subtype was enriched for APOE ε3 (85.7%) and had the lowest intermediate-to-high ADNC (2.4%). The “pTau” subtype showed moderate dementia prevalence (58.9%) and increased intermediate-to-high ADNC frequency (44.6%), whereas the “α-synuclein 2” subtype had rare intermediate-to-high ADNC (7.1%). Male sex was slightly more frequent in subtypes 4 (71.4%) and 3 (66.1%) than in subtypes 2 (62.0%) or 1 (56.0%). Shorter disease duration (<15 years) was more common in subtype 3 (57.1%), while longer duration (≥15 years) was more frequent in subtype 2 (57.8%). Dementia prevalence, ischaemic pathology, OH, and age at death did not differ significantly across subtypes. Pairwise comparisons using Chi-square tests confirmed significant differences in APOE genotype (adj. p = 0.04999) and AD co-pathology (adj. p = 0.00048) between “Aβ” and “α-synuclein 1”, and in AD co-pathology between “α-synuclein 1” and “pTau” (adj. p < 0.000001) (Supplementary Data for Fig. 7).

Pathology burden differentiated the SuStaIn subtypes. α-Synuclein load was higher in the “α-synuclein 1” subtype than in the “Aβ” and “pTau” subtypes. Aβ burden was greatest in the “Aβ” subtype and higher in “pTau” than in “α-synuclein 1”. pTau burden was selectively elevated in the “pTau” subtype. These results indicate that SuStaIn-defined subtypes are distinguished mainly by dominant misfolded protein profiles rather than demographic or clinical features (Supplementary Fig. 5, Supplementary Data).

## Discussion

Our findings redefine clinicopathological associations in primary LBD, highlighting genotype-specific dementia risk and revealing novel progression patterns through data-driven modelling.

### Quantitative pathology outperforms traditional staging in predicting dementia risk

Our results highlight advantages of automated quantitative pathology assessment over conventional staging. Whereas traditional schemes capture only anatomical spread, quantitative thresholds demonstrate intensifying pathology within affected regions, correlate more strongly with dementia, enable finer genotype-pathology integration, and provide reproducible digital standardisation. We show that LBD progression reflects both the spatial spread of Lewy, Aβ, and pTau pathology and rising regional burden, with particularly marked amygdala Lewy accumulation, suggesting AD co-pathology influences Lewy pathology deposition in this region. For dementia prediction, cortical Lewy pathology burden was a stronger predictor than regional distribution alone, outperforming Braak staging (2003),^22^ and given their structural similarity, is also expected to outperform other schemes such as the unified staging system (2009),^39^ McKeith criteria (2017) and neuropathological consensus criteria (2021).^39^ While cortical Lewy pathology burden has been linked to cognitive impairment in PD using manual counts,^5,40^ our pipeline enables large-scale automated assessment.

These findings emphasise that incorporating regional burden metrics into staging frameworks may improve clinicopathological correlation, prediction of disease milestones, and understanding of disease progression.

TDP-43 pathology was rare and correlated with age as reported in the general population,^41^, suggesting it is not a major dementia risk factor in our LBD cohort.

### Male sex increases dementia risk in specific clinicopathological contexts

Biological sex influenced dementia probability when evaluated with age at death, disease duration and ischaemic pathology. Male sex increased risk in both APOE genotypes, with a stronger effect in ε3 carriers, particularly with ischaemic pathology present, suggesting vascular mechanisms may underlie this subgroup effect. Although APOE ε4 remained the main dementia risk factor, male sex further increased risk in ε4 carriers with longer disease duration and older age at death. These findings are consistent with reports of higher vascular comorbidity in men with dementia,^42^ although the rate of age-related cognitive decline is reported to be faster in females.^43^ This suggests that sex-specific dementia risk modulation in LBD may depend on both genotype and co-pathology. Future studies should investigate vascular, hormonal, and inflammatory mechanisms underlying these associations and test whether sex-stratified risk models improve dementia prediction.

### APOE ε4 impacts regional pathology burden and dementia risk in LBD

APOE ε4 carriers showed higher cortical and limbic, but not brainstem, Lewy pathology. SuStaIn modelling identified an ε4-enriched subgroup with early amygdala involvement and milder, concomitant brainstem pathology. Comparable ε4 frequency in PD and PDD, but regional Lewy burden differences in dementia, suggest ε4 modulates progression rather than onset, consistent with reports linking ε4 to greater cortical α-synuclein, Aβ, and dementia risk in LBD.^8–10^ PD ε4 carriers without dementia had Lewy, Aβ and pTau burdens comparable to PDD ε3 carriers, consistent with ε4’s established role in accelerating Aβ deposition in AD,^44–46^ and suggesting a similar mechanism in LBD. Potential pathways include impaired Aβ clearance, enhanced seeding, and altered lipid metabolism that may facilitate Aβ and α-synuclein aggregation.^47,48^ Experimental studies further show that APOE ε4 can exacerbate α-synuclein pathology and related neurotoxicity independently of Aβ^49^ and amplify α-synuclein seeding activity in AD with Lewy pathology,^50^.

Quantitative burden analyses indicate that dementia risk in LBD is shaped by genotype-specific thresholds and by interactions between α-synuclein and AD co-pathology. Across both APOE ε3 and ε4 carriers, α-synuclein burden was the principal determinant of risk, but ε3 carriers appeared more sensitive to its accumulation. Our post-mortem analyses showed that ε4 carriers with dementia had higher Lewy pathology and Aβ burden than ε3 carriers, supporting the role of APOE ε4 in conferring dementia risk. These findings suggest that ε4 both promotes greater accumulation of pathology and modifies the threshold for dementia onset, with ε4 carriers tolerating higher α-synuclein and Aβ load before dementia onset. Longitudinal quantitative in-vivo imaging with clinical and genotype correlation will be required to test this directly. The absence of independent predictive value for pTau indicates it is largely downstream of Aβ, consistent with the amyloid cascade hypothesis.^51^

Taken together, these findings support a multifactorial role for APOE ε4 in both α-synuclein and Aβ accumulation, whereby enhanced amyloidogenic processes interact with α-synuclein to shape regional, particularly limbic, vulnerability and divergence in Lewy pathology progression patterns.

### Protein burden and APOE genotype modulate ischaemic effects in LBD

Previous work has linked OH to cognitive decline in PD likely via hypoperfusion.^16,17^ In our cohort, OH frequency was similar in demented and non-demented LBD cases, but when combined with ischaemic pathology it amplified dementia risk in cases with low misfolded protein burden. At high burden, this effect disappeared, suggesting proteinopathy overrides vascular modifiers.

### Data-driven modelling identifies divergent Lewy body progression patterns

Earlier microarray studies of α-synuclein, Aβ, and pTau, in 159 post-mortem cases (controls, AD, LBD, and mixed), identified clinically relevant α-synuclein topographies.^52^ Our study is the first large-scale analysis of primary LBD using automated quantification and progression modelling. SuStaIn modelling confirmed the canonical caudal-to-rostral spread of Lewy pathology,^22^ but also showed marked heterogeneity, consistent with earlier work, including neuroimaging SuStaIn studies in PD^53^ and DLB,^54^ and regional post-mortem α-synuclein pathology in population cohort with Lewy pathology,^55^ highlighting the value of data-driven pathological subtyping in capturing biologically and clinically relevant variability.

Alongside the conventional brainstem-first trajectory, we identified a subtype with early amygdala involvement concomitant with brainstem pathology. In line with previous reports,^55,56^ this subtype was enriched for APOE ε4 and advanced AD co-pathology, whereas the canonical brainstem subtype was enriched for APOE ε3 carriers, and had lower AD co-pathology and dementia prevalence. Presence of ε4 carriers in the brainstem subtype, and ε3 carriers in the amygdala subtype, suggests that additional genetic factors or local microenvironmental cues may lead to selective regional vulnerability to Lewy pathology.

We found no evidence that Lewy pathology originates solely within the amygdala with secondary spread to brainstem or cortical regions. Our findings show early amygdala involvement occurring in parallel with brainstem pathology, contrasting with findings from population-based studies^55–57^ and observations in AD,^11^ where the amygdala can be the only region affected by Lewy pathology. Biologically, this suggests that the amygdala is unlikely to serve as an independent nidus of α-synuclein pathology in primary LBD; rather, its early involvement in a subset of patients reflects local environment and genotype-driven selective vulnerability. APOE ε4 is a known risk factor for earlier Aβ pathology formation and altered lipid metabolism,^58^ which has been linked to α-synuclein aggregation and toxicity in experimental systems.^49,50,59^ Moreover, aggregated protein-lipid compartmentalisation with organellar crowding has been demonstrated in Lewy bodies from post-mortem PD brains,^60^ indicating that APOE ε4-related lipid metabolism impairment may promote α-synuclein aggregation in vulnerable regions. In AD and other neurodegenerative diseases, isolated amygdala Lewy pathology likely reflects a genotype-associated process distinct from canonical LBD propagation.

We also identified two novel Lewy pathology propagation patterns. One showed early anterior cingulate involvement occurring alongside brainstem pathology. The other subtype showed early isolated neocortical involvement and more frequent OH. The neocortical subtype was enriched for APOE ε4 and advanced AD co-pathology, like the amygdala subtype. Both, the neocortical and early amygdala subtypes had higher dementia prevalence even without AD co-pathology, supporting ours and previous findings^5,40^ that Lewy pathology burden is an independent dementia risk factor, highlighting the value of quantitative assessment for identifying high-risk individuals. Notably, neither of these two novel subtypes was detected in the previous SuStaIn study of more than 800 cases from a mixed population,^55^ suggesting they represent distinct, novel progression patterns in primary LBD.

Multimodal modelling of α-synuclein, Aβ, and pTau revealed additional spatiotemporal patterns. LBD cases from our cohort could be classified by whether Aβ or pTau accumulation preceded Lewy pathology. The identification of Aβ-first and AD-type pTau-first trajectories supports the hypothesis that, in some patients, AD-type co-pathologies precede and potentially promote α-synuclein aggregation. In contrast, subtype 2, a canonical caudal/limbic-to rostral trajectory, showed the lowest AD co-pathology burden and was enriched for APOE ε3 carriers, suggesting a predominantly α-synuclein-driven process. Another atypical α-synuclein subtype exhibited early parietal cortex involvement, a region typically affected late in PD. Notably this atypical Lewy pathology progression subtype was enriched for cases with none to low levels of concomitant AD pathology, yet half of the cases had clinical evidence of dementia, adding to the evidence that cortical α-synuclein deposition alone is sufficient to drive cognitive decline in some individuals.

Together, these findings indicate that misfolded protein propagation in primary LBD follows distinct trajectories, linked to genotype, co-pathology, and clinical features. APOE associations suggest genotype-specific regional and likely cellular vulnerabilities, while the identification of atypical cortical-dominant patterns challenges existing pathology staging models. This highlights the importance of subtype-specific approaches to diagnosis and incorporating both genetic risk and co-pathology status into protein-based disease modifying trial designs.

The main strength of this study is the validation and integration of automated quantitative measurements of α-synuclein, Aβ, and AD-type pTau pathology across multiple brain regions from WSI, with the APOE genotype, and clinical variables in a primary LBD cohort.

Our findings should be interpreted in the context of several limitations. Clinical information on OH and dementia may be incomplete owing to retrospective reporting. APOE genotype data were unavailable for some cases, limiting the ability to fully assess genotype-specific effects. Histological examination was restricted to one hemisphere, which may underestimate microscopic ischaemic burden if pathology was confined to the contralateral side. We also did not assess the extent or severity of large- and small-vessel disease. Furthermore, the absence of olfactory bulb tissue in our cohort precluded direct comparison with an earlier study identifying it as a frequent initial site of Lewy pathology.^55^ Similarly, the absence of peripheral autonomic tissues precluded comparison with recent study that identified vagal and sympathetic body-first subtypes of LBD.^61^ Finally, modest sample sizes in genotype-stratified analyses and the cross-sectional design make it difficult to determine whether the observed associations are causal.

Despite these limitations, our study has significant translational value. The automated pipeline we developed for large-scale quantification of misfolded proteinopathies provides a foundation for standardising digital pathology data across neurodegenerative diseases. The distinct Lewy pathology propagation patterns we have identified by integrating APOE genotype, AD co-pathology, ischaemic pathology, and clinical features, offer a framework for improved clinical risk stratification and targeted therapies and highlight the need for multidimensional risk models.

We show that APOE ε3 and ε4 carriers differ in their pathological thresholds for dementia, generating testable hypotheses for genotype-tailored therapies and in defining thresholds for protein-based biomarkers or PET-imaging measures. Our findings may aid interpretation of emerging seeding aggregation assays for fluid-based detection of α-synuclein, Aβ, and pTau biomarkers, and support comprehensive GWAS analyses within identified patient subgroups to uncover additional genetic risk factors for dementia and other clinical milestones. Validation of our findings in independent longitudinal cohorts will be crucial.

In conclusion, by integrating quantitative misfolded protein burden with genotype and clinical data, we define biologically distinct but overlapping pathways to dementia in primary LBD. These results refine current models of disease progression, provide mechanistic hypotheses for genotype- and pathology-specific therapies, and establish a quantitative digital pathology based-framework for future biomarker development and disease-modifying therapy trials.

## Data availability

The raw quantitative pathological data, protocols, R and python code and key lab materials used and generated in this study are listed in a Key Resource Table (KRT) alongside their persistent identifiers at 10.5281/zenodo.16994814. Whole slide images of cases used in this study are available for viewing via UCL QSBB Digital Pathology Resource (RRID:SCR_025020). Anonymised clinical and pathology data are available upon reasonable request, if in compliance with regulatory ethical approval.

## Acknowledgements

We thank the patients and their relatives for their invaluable contribution through brain donation. We thank UCL QSBB administrative and technical staff for their assistance. We thank Oke Avwenagha for her assistance. All genotyping data used in the preparation of this article were obtained from Global Parkinson’s Genetics Program (GP2). GP2 is funded by the Aligning Science Across Parkinson’s (ASAP) initiative and implemented by The Michael J. Fox Foundation for Parkinson’s Research (https://gp2.org). For a complete list of GP2 members see https://gp2.org. A large language model (ChatGPT, OpenAI) was used to improve the clarity and conciseness of specific phrases and sentences in the manuscript. Figure 1 was created using BioRender (https://www.biorender.com) and Figure 5C was created using draw.io (https://www.drawio.com).

## Supplementary material

The following supplementary material are available at *Brain* online: Supplementary methods and supplementary figure legends.

Supplementary Figure 1: Influence of dementia status on regional α-synuclein, Aβ and pTau pathology in Lewy body disease stratified by APOE genotype.

Supplementary Figure 2: SuStaIn modelling of regional Lewy pathology burden.

Supplementary Figure 3: Mean regional Lewy, Aβ, and pTau pathology across SuStaIn subtypes of Lewy pathology progression.

Supplementary Figure 4: SuStaIn modelling of combined Lewy, Aβ, and pTau pathology.

Supplementary Figure 5: Demographics and regional pathology distribution across SuStaIn subtypes of combined Lewy, Aβ, and pTau progression.

Supplementary Table 1: Abbreviations for regions of interest.

Supplementary Table 2: Variables included in radar plots (Figure 1).

Supplementary Table 3: Clinical and demographic characteristics of Lewy body disease cohorts stratified by APOE status.

Supplementary Table 4: Clinical and demographic characteristics of Lewy body disease cohorts stratified by dementia and APOE status.

Supplementary Table 5: Clinical and demographic characteristics of Lewy body disease cohorts stratified by TDP-43 LATE stage.

Supplementary Table 6. Measurements collected for each pseudo-cell in Lewy body classifier. Supplementary Excel statistics data for Figures.

Supplementary Excel statistics data for Supplementary Figures. Supplementary Excel statistics data for Tables.

Supplementary Excel statistics data for Supplementary Tables.

