## Supplementary data and figures for "Quantitative pathology and APOE genotype reveal dementia risk and progression in Lewy body disease"

**Affiliations**

### Supplementary methods

#### Immunohistochemical staining

7 µm sections were cut from formalin-fixed, paraffin-embedded tissue blocks from representative brain regions and mounted on glass slides (see Supplementary Table 1 for a complete list of regions). Immunohistochemical staining for α-synuclein (MA1-90342; Thermo Scientific; 1:1500), amyloid-β (M0872; Dako; 1:100), phosphorylated tau (MN1020; AT8; Thermo Scientific; 1:1,200) and non-phosphorylated TDP-43 (2E2-D3, H00023435-M01, Abnova, 1:500) was performed using automated staining platforms from Menarini Diagnostics or Ventana (Roche). Biotinylated secondary antibodies were used with horseradish peroxidase-conjugated streptavidin complex and diaminobenzidine as the chromogen, with haematoxylin counterstaining. Appropriate positive and negative controls were included in each staining run. Slides were also stained for haematoxylin & eosin. Pathological diagnoses were confirmed by neuropathologists experienced in neurodegenerative diseases using relevant consensus diagnostic and staging criteria.[1-7]

#### Quantitative digital neuropathology

The following image analysis methods were performed in QuPath v0.5.1 after importing annotations.[8]

##### a. Colour deconvolution and background correction:

To address batch effects in stain intensity, stain vectors for colour deconvolution were calculated from a composite image consisting of 1000×1000 pixels squares derived from a subset of 270 whole slide images (WSI), randomly selected from across the entire cohort and stained for α-synuclein, amyloid-β (Aβ) and phosphorylated tau (pTau), with 1% of extreme pixels ignored. This is to account for variability in haematoxylin and DAB staining intensities across the years that the slides were stained and to establish stain vectors that are specific for the colour profile of the slide scanners used in this study. After applying the stain vectors onto each WSI, to further address batch variations in staining intensity, the mean deconvoluted DAB intensity within each "!BACKGROUND" (BG) annotation was calculated (DAB: Mean (BG)) and saved as a measurement in their corresponding brain region annotation. The BG annotation is described in the main Methods. The resulting DAB-channel specific images were then analysed for Aβ and pTau using automatic thresholding and for Lewy bodies using the deep learning-based segmentation algorithm *StarDist,*[9] respectively.

##### b. Aβ and pTau automatic thresholding:

For each brain region, a threshold for positive DAB staining was calculated using automatic thresholding methods Triangle[10] or Otsu[11] via an adaptation of FIJI/ImageJ's implementation (https://github.com/fiji/Auto_Threshold). Using the calculated background DAB intensity and the automatic threshold values for positive DAB staining, the following threshold values were derived:

$$Background\ corrected\ fixed\ threshold\ value = DAB\!:Mean\ (BG) + 0.25$$

$$Background\ corrected\ Triangle\ threshold\ value\ (Amyloid\ \beta) \\ = DAB\!:Mean\ (BG) + Triangle\ threshold\ value$$

$$Background\ corrected\ Otsu\ threshold\ value\ (pTau) \\ = DAB\!:Mean\ (BG) + Otsu\ threshold\ value$$

The derived background-corrected threshold values were then used to measure the area of positive DAB staining, Triangle[10] for Aβ and Otsu[11] for pTau, respectively, within each brain region annotation. Finally, the percentage of positive DAB staining within each brain region was calculated by:

$$\%\,positive\ DAB\ staining\ area = \frac{Area\ of\ positive\ DAB\ staining}{Area\ of\ brain\ region\ annotation} \times 100$$

These values were then plotted as either % area of Aβ (abbreviated as %Aβ in Figures and Tables) or % area of phosphorylated tau (abbreviated as %pTau in Figures and Tables), respectively.

##### c. Lewy body classifier:

α-synuclein images were pre-processed similarly by importing WSI and region of interest annotations and then applying stain vectors for colour deconvolution. To detect Lewy bodies, the deep learning-based segmentation algorithm *StarDist*[9] was used on the deconvoluted DAB channel. The resulting detections were dilated by 2 µm to create a pseudo-cell where the "nucleus" is the original detection and the "cytoplasm" is the area surrounding the "nucleus" after 2 µm dilation. This is used to measure the background DAB intensity surrounding each detection object. For each pseudo-cell, the following shape and intensity measurements were measured: intensity, shape/morphology and Haralick as described in Supplementary Table 6. These measurements were used to train a random trees object classifier from a set of 40 training images to classify Lewy bodies from non-specific detections. The resulting Lewy body object classifier was then applied to the entire cohort of WSIs. The resulting Lewy body detections were then divided by the area to give measurements of Lewy bodies per $mm^2$ for each region of interest (abbreviated as LB/$mm^2$ in Figures and Tables).

### Uni- and multivariate associations with dementia risk

#### a. Cumulative risk heatmap

To evaluate dementia risk across all combinations of key pathology and demographic predictors, we constructed a cumulative risk matrix using an exhaustive subgrouping approach based on seven binary variables: α-synuclein burden (high vs low, 75$^{th}$ percentile cutoff), Aβ burden (high vs low, 75$^{th}$ percentile cutoff), pTau burden (high vs low, 85$^{th}$ percentile cutoff), ischaemic pathology (present vs absent), biological sex (male vs female), disease duration (<15 vs ≥15 years), and age at death (<77 vs ≥77 years). These groupings were generated separately for APOE ε3 and ε4 carriers.
All 128 possible combinations of the seven binary variables were constructed using the expand.grid() function in R.[12] The resulting predicted probabilities were visualized in a heatmap matrix to illustrate how dementia risk varies across strata of pathology burden and demographic, and clinical parameters.

#### b. Rule-based surrogate decision tree

To enable interpretable subgroup-level stratification, a rule-based surrogate decision tree was constructed using seven binary predictors. APOE ε3 and ε4 groups were analysed separately, with cutoffs for α-synuclein and Aβ set at the 75$^{th}$ percentile and for pTau at the 85$^{th}$ percentile, calculated within each genotype group. For each of the 128 possible combinations of binary predictors, dementia probability was estimated using predictions from the logistic regression model. Variables were arranged in the tree according to decreasing magnitude of genotype-specific logistic regression model coefficients (odds ratios) for dementia risk, with the strongest predictors placed nearest the root. We evaluated model performance using ROC analysis with the pROC package in R,[13] reporting the area under the ROC curve (AUC) and the optimal probability threshold determined by Youden's J statistic,[14] together with the corresponding sensitivity and specificity values. All evaluations were performed separately within each genotype subgroup using stratified 10-fold cross-validation.

#### Regional quantitative pathology-driven disease progression modelling

SuStaIn simultaneously infers subtype-specific progression sequences and probabilistic subtype/stage assignments from cross-sectional data using a generative modelling framework.[15] Model uncertainty was assessed with 5,000,000 Markov Chain Monte Carlo (MCMC) iterations, and the expectation-maximization process optimised the subtype sequences from 30 random starting points to find the maximum likelihood solution. Model selection was guided by 10-fold cross-validation using the cross-validation information criterion (CVIC) at 5,000,000 MCMC iterations, with lower values indicating better fit, tested from 1-7 subtypes. We also examined test-set log-likelihoods to evaluate generalizability, and MCMC trace plots and histograms to confirm stability across subtypes. The optimal Lewy pathology model and combined Lewy, Aβ, and pTau pathology model supported 4 subtypes as, despite decreasing CVIC values with increasing complexity, there was little increase in test-set log-likelihoods (Supplementary Figs 2 and 4, Supplementary Data).[16] Each SuStaIn subtype in Figs 6 and 7 are visualized using a positional variance diagram, which illustrates the most probable sequence of regional involvement across SuStaIn stages, along with the associated uncertainty. Regional pathology values were standardised against the synthetic control distribution, such that the control mean defines the baseline (0) and each unit reflects one control standard deviation (SD). In the SuStaIn positional variance diagrams, three abnormality thresholds relative to controls are shown: mild (≥1 SD, red), moderate (≥2 SD, magenta), and severe (≥3 SD, blue). Colour intensity indicates the certainty with which the model assigns a biomarker to a given abnormality threshold at each disease stage: deeper hues denote higher certainty, paler hues indicate greater positional variance (uncertainty), and white reflect minimal or no probability.

#### Statistical analysis

For pathology burden classification and diagnostic performance metrics, quantitative measures of each misfolded proteinopathy (α-synuclein, Aβ, and pTau) were compared between binarised subgroups using the Kruskal–Wallis test followed by Dunn's pairwise post-hoc comparisons with Benjamini–Hochberg (BH)[17] p-value correction. Spearman's rank correlation was used to evaluate the association between semi-quantitative pathology scores and their corresponding quantitative burdens (Spearman ρ: α-synuclein = 0.72, Aβ = 0.80, pTau = 0.76; Spearman $p < 0.0001$ for all three pathologies), as visualized in scatter plots in Fig. 1. Kruskal-Wallis test with Dunn-BH correction was employed to compare: (i) traditional pathology staging schemes vs. quantitative misfolded protein pathological burdens (Fig. 1) and (ii) regional pathology across quantitative misfolded protein pathologies (Figs 2A, 2B, 3, Supplementary Figs 3 and 5, Supplementary Data).

Two-sided Kolmogorov-Smirnov tests were used to compare the distribution of quantitative pathology burden between APOE ε3 and ε4 carriers (Fig. 2C, Supplementary Data). Chi-square or Fisher's exact tests (when expected counts were <5) were used to compare proportions of dementia cases across semi-quantitatively determined pathology stages (Braak stages for Lewy pathology, Thal phases for Aβ and Braak & Braak stages for pTau) and quantitative pathological groupings (Fig. 2D, Supplementary Data) and all categorical clinical and demographic variables (Table 1, Supplementary Tables 3, 4 and 5, Sankey diagrams for Figs 6 and 7, and Supplementary Data). Mann-Whitney U tests with BH correction were used to compare regional quantitative Lewy, Aβ and pTau pathology between APOE ε3 and ε4 genotypes (Fig. 3, Supplementary Data) and between PD cases with and without dementia (Supplementary Fig. 1). A binary univariate logistic regression model was fitted for the presence and absence of ischaemic pathology, separately for cases with low and high misfolded protein pathology burden respectively (Fig. 4B) and a multivariate generalized linear model (glm) was fitted separately for APOE ε3 and ε4 carriers for the following variables: α-synuclein burden, Aβ burden, pTau burden, ischaemic pathology (present vs absent), biological sex (male vs female), disease duration (<15 vs ≥15 years), and age at death (<77 vs ≥77 years) and significance was assessed using Wald tests on the model coefficient for each variable (Fig. 5A and Supplementary Data). For both regression analyses, the outcome variable was the presence or absence of dementia.

### Supplementary figures and figure legends:

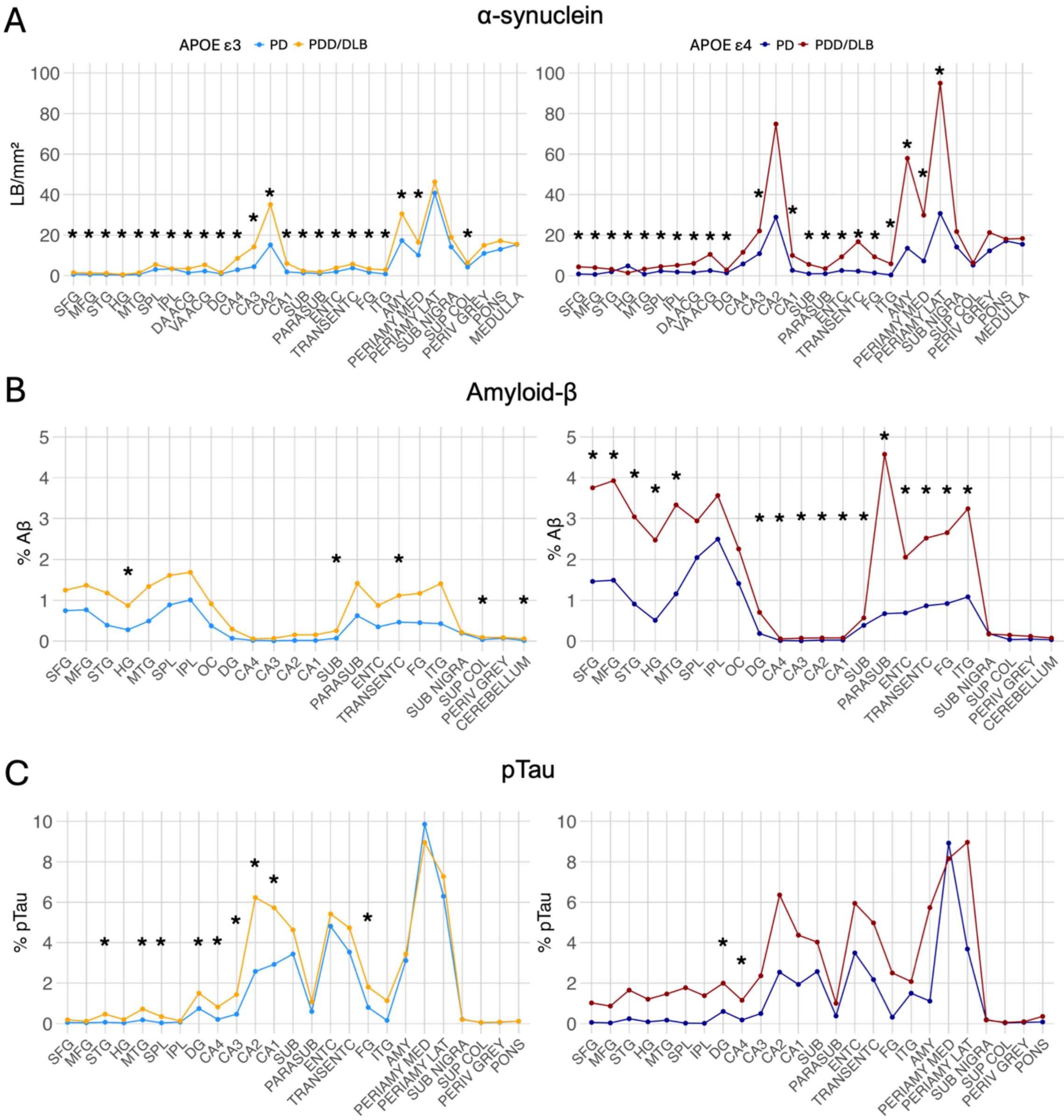


**Supplementary Figure 1: Influence of dementia status on regional α-synuclein, Aβ and pTau pathology in Lewy body disease stratified by APOE genotype.**

**(A)** Lewy body (LB/mm$^2$), **(B)** % area of Aβ (%Aβ) and **(C)** % area of phosphorylated tau (%pTau) quantitative pathology burden across multiple brain regions in APOE ε3 and ε4 carriers, stratified by dementia status (light blue APOE ε3 PD, yellow APOE ε3 PDD/DLB, dark blue APOE ε4 PD, brown APOE ε4 PDD/DLB).

Lines indicate group mean pathology burden across regions; * indicates statistically significant differences between dementia (PD vs PDD/DLB) groups in each region ($p < 0.05$; statistical test Mann-Whitney test with BH correction per region for each proteinopathy). Brain region abbreviations are detailed in Supplementary Table 1 and detailed statistical outputs are available in supplementary data (Excel statistics) for Supplementary Figure 1.

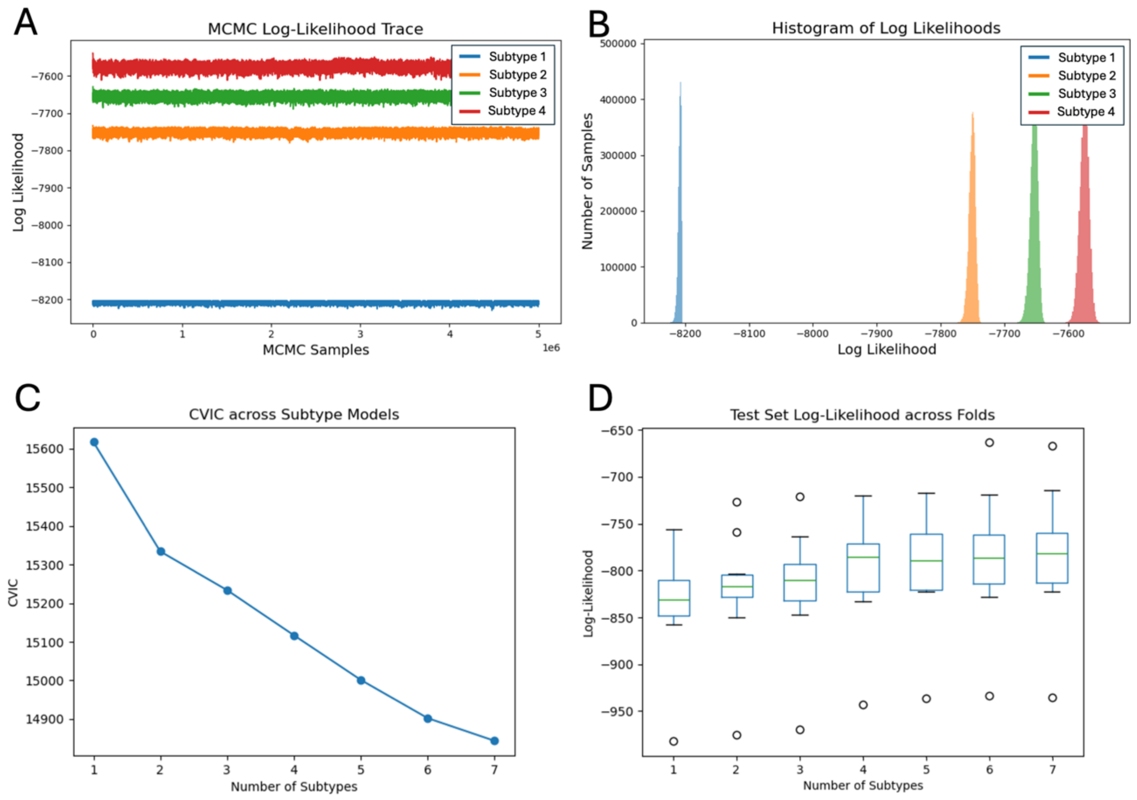


**Supplementary Figure 2: SuStaIn modelling of regional Lewy pathology burden.**

**(A)** Markov Chain Monte Carlo (MCMC) log-likelihood traces for four subtypes, showing stable convergence across 5 million iterations based on regional Lewy pathology. **(B)** Histograms of log-likelihood values for the four subtypes, illustrating the distribution across MCMC iterations. **(C)** Ten-fold cross-validation information criterion (CVIC) values for models with one to seven subtypes, with lower CVIC values indicating better fit. **(D)** Boxplots of test-set log-likelihood across ten cross-validation folds for one to seven subtype models, demonstrating model generalisability. Detailed outputs are available in supplementary data (Excel statistics) for Supplementary Figure 2.

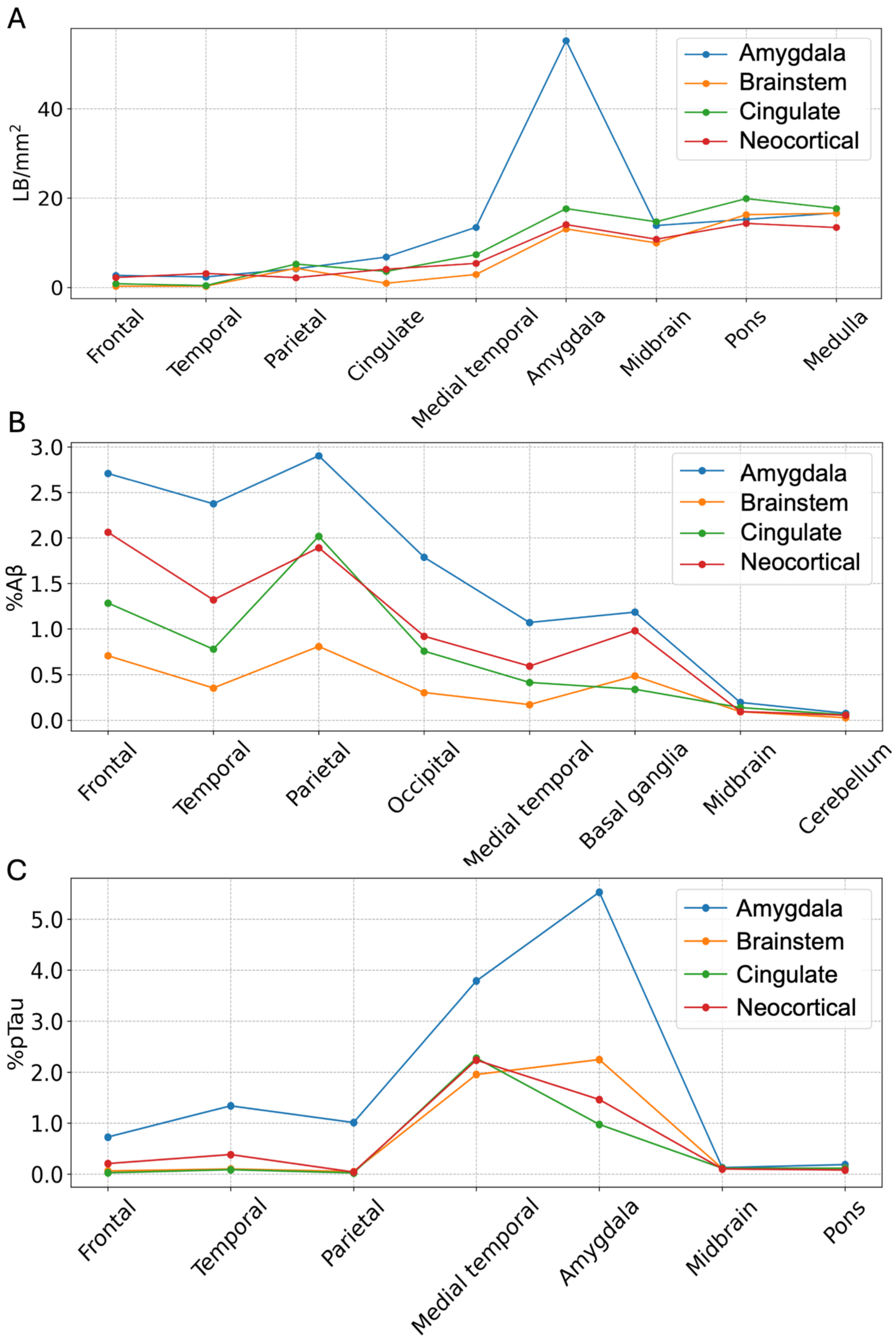
A
LB/mm$^2$
0
20
40
Amygdala
Brainstem
Cingulate
Neocortical
Frontal
Temporal
Parietal
Cingulate
Medial temporal
Amygdala
Midbrain
Pons
Medulla
B
%Aβ
0.0
0.5
1.0
1.5
2.0
2.5
3.0
Amygdala
Brainstem
Cingulate
Neocortical
Frontal
Temporal
Parietal
Occipital
Medial temporal
Basal ganglia
Midbrain
Cerebellum
C
%pTau
0.0
1.0
2.0
3.0
4.0
5.0
Amygdala
Brainstem
Cingulate
Neocortical
Frontal
Temporal
Parietal
Medial temporal
Amygdala
Midbrain
Pons

**Supplementary Figure 3: Mean regional Lewy, Aβ, and pTau pathology across SuStaIn subtypes of Lewy pathology progression.**

Each line shows mean quantitative **(A)** Lewy body (LB/$mm^2$), **(B)** % area of Aβ (%Aβ) and **(C)** % area of phosphorylated tau (%pTau) pathology burden across multiple brain regions, stratified by SuStaIn-inferred subtypes of Lewy pathology progression based on quantitative Lewy body burden. Each line represents the mean pathology burden for a subtype across regions. Brain regions include average quantitative scores from frontal (cortices of the superior frontal and middle frontal gyri), temporal (cortices of the superior temporal, Heschl's and middle temporal gyri), parietal (cortices of the superior parietal and inferior parietal lobules), occipital (occipital cortex), medial temporal (dentate gyrus, CA4, CA3, CA2, CA1, subiculum, parasubiculum, entorhinal cortex, transentorhinal cortex, cortex of fusiform gyrus), amygdala, basal ganglia (caudate nucleus and putamen) midbrain (substantia nigra, superior colliculus, periventricular grey matter), pons (pontine tegmentum), medulla (medullary tegmentum) and cerebellum. Detailed statistical outputs for Kruskal-Wallis test with Dunn's pairwise post-hoc comparisons and Benjamini-Hochberg (BH) p-value adjustment in supplementary data (Excel statistics) for Supplementary Figure 3.

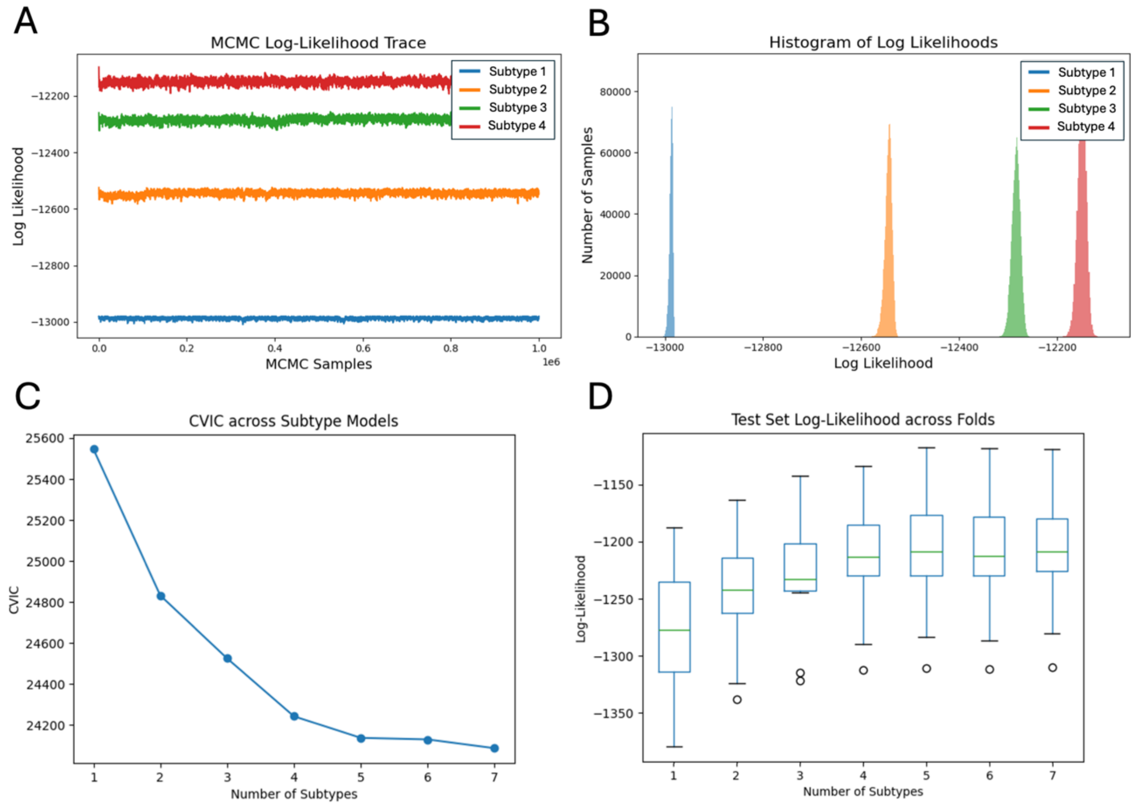


**Supplementary Figure 4: SuStaIn modelling of combined Lewy, Aβ, and pTau pathology.**

**(A)** Markov Chain Monte Carlo (MCMC) log-likelihood traces for four subtypes based on frontal cortical and medial temporal lobe quantitative Lewy, Aβ, and pTau pathology, showing stable convergence across 5 million samples. Cortical regions include cortices of the superior and middle frontal gyri, cortices of the superior and middle temporal and Heschl's gyri, and cortices of the superior and inferior parietal lobules; medial temporal lobe regions include dentate gyrus, CA4, CA3, CA2, CA1, subiculum, parasubiculum, entorhinal cortex, transentorhinal cortex and cortex of the fusiform gyrus. **(B)** Histograms of log-likelihood values for the four subtypes, illustrating the distribution across MCMC iterations. **(C)** Ten-fold cross-validation information criterion (CVIC) values for models with one to seven subtypes, with lower CVIC values indicating better fit. **(D)** Boxplots of test-set log-likelihood across ten cross-validation folds for one to seven subtype models, demonstrating model generalisability. Detailed outputs are available in supplementary data (Excel statistics) for Supplementary Figure 4.

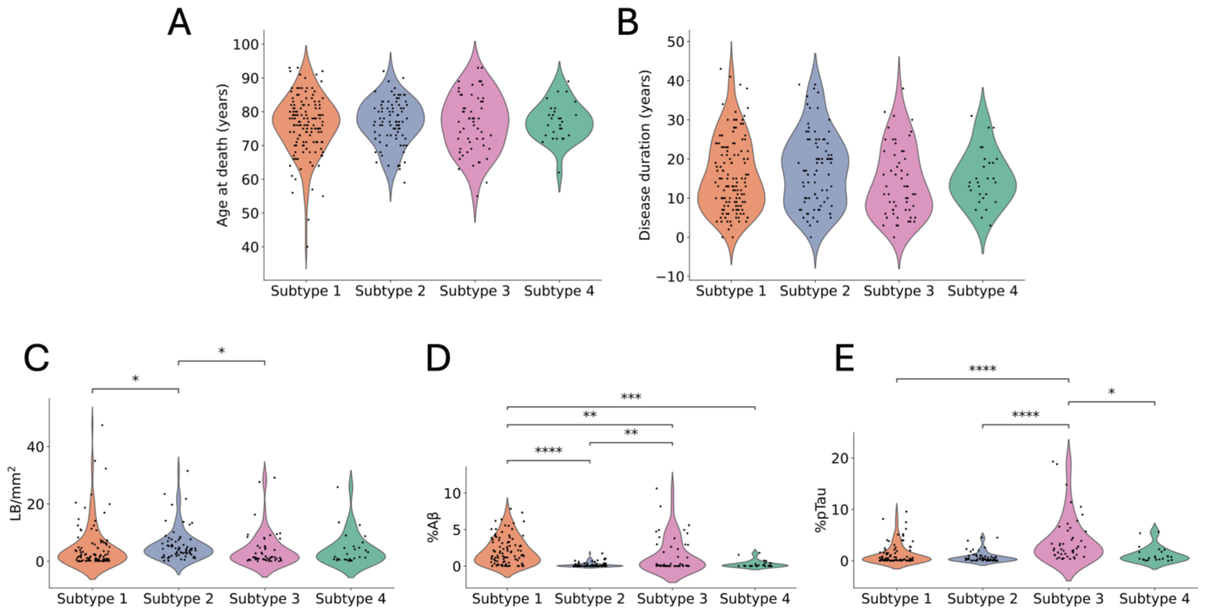


**Supplementary Figure 5: Demographics and regional pathology distribution across SuStaIn subtypes of combined Lewy, Aβ, and pTau progression.**

**(A)** Age at death (years) and **(B)** disease duration (years) for SuStaIn-inferred spatiotemporal trajectories. **(C)** Lewy body (LB/mm$^2$), **(D)** % area of Aβ (%Aβ) and **(E)** % area of phosphorylated tau (%pTau) quantitative pathology burden across frontal cortical and medial temporal lobe regions, stratified by SuStaIn-inferred spatiotemporal progression subtypes of combined Lewy body density (LB/mm$^2$), %Aβ and %pTau pathology. Frontal regions include cortices of the superior and middle frontal gyri and medial temporal regions include dentate gyrus, CA4, CA3, CA2, CA1, subiculum, parasubiculum, entorhinal cortex, transentorhinal cortex and cortex of the fusiform gyrus. Kruskal-Wallis test with Dunn's pairwise post-hoc comparisons and Benjamini-Hochberg (BH) p-value adjustment, (* $p \leq 0.05$, ** $p \leq 0.01$, *** $p \leq 0.001$, **** $p \leq 0.0001$). Detailed outputs are available in supplementary data (Excel statistics) for Supplementary Figure 5.

### Supplementary tables:

**Supplementary Table 1: Abbreviations for regions of interest.**

| Region abbreviation | Anatomical region |
|---|---|
| **Frontal Cortex** | |
| SFG | Superior Frontal Gyrus (Grey Matter) |
| MFG | Middle Frontal Gyrus (Grey Matter) |
| **Temporal Cortex** | |
| STG | Superior Temporal Gyrus (Grey Matter) |
| MTG | Middle Temporal Gyrus (Grey Matter) |
| HG | Heschl's Gyrus (Grey Matter) |
| **Parietal Cortex** | |
| SPL | Superior Parietal Lobule (Grey Matter) |
| IPL | Inferior Parietal Lobule (Grey Matter) |
| **Occipital Cortex** | |
| OC | Occipital (Grey Matter) |
| **Cingulate Cortex** | |
| VA ACG | Ventro-Anterior Cingulate Cortex (Grey Matter) |
| DA ACG | Dorso-Anterior Cingulate Cortex (Grey Matter) |
| **Hippocampus** | |
| DG | Dentate Gyrus |
| CA4 | CA4 |
| CA3 | CA3 |
| CA2 | CA2 |
| CA1 | CA1 |
| SUB | Subiculum |
| PARASUB | Parasubiculum |
| ENTC | Entorhinal Cortex |
| TRANSENTC | Transentorhinal Cortex |
| FG | Fusiform Gyrus |
| ITG | Inferior Temporal Gyrus |
| **Amygdala** | |
| AMY | Amygdala |
| AMY_LS | Amygdala Lateral |
| AMY_MI | Amygdala Medial |
| PERIAM MED | Periamygdaloid Cortex Medial |
| PERIAM LAT | Periamygdaloid Cortex Lateral |
| **Midbrain** | |

| | |
|---|---|
| PERIV GREY | Periventricular Grey Matter |
| SUP COL | Superior Colliculus |
| SUB NIGRA | Substantia Nigra |
| **Pons** | |
| PONS | Pontine Tegmentum |
| **Medulla** | |
| MEDULLA | Medulla Tegmentum |
| **Cerebellum** | |
| CC | Cerebellar Cortex |

List of abbreviations used to denote specific neuroanatomical regions of interest that are analysed for regional semi-quantitative and quantitative digital pathology.

**Supplementary Table 2: Variables included in radar plots (Figure 1).**

| **Metric** | **Equation** | **Description** |
|---|---|---|
| Sensitivity | Sensitivity = TP / (TP + FN) | Proportion of dementia cases with high pathology correctly identified (true positive rate) |
| Specificity | Specificity = TN / (TN + FP) | Proportion of non-dementia cases with low pathology correctly identified (true negative rate) |
| Odds Ratio (OR) | OR = (TP × TN) / (FP × FN) | Measure of association between dementia status and pathology burden |
| Positive Predictive Value (PPV) | PPV = TP / (TP + FP) | Probability of dementia when pathology is high |
| Pathology Absence Ratio (PAR) | PAR = Dementia Low Path / Total Low Path | Proportion of dementia cases when pathology is low |
| Pathology Presence Ratio (PPR) | PPR = Dementia High Path / Total High Path | Proportion of dementia cases when pathology is high |

TP = True Positives (dementia & high pathology), TN = True Negatives (no dementia & low pathology), FP = False Positives (no dementia & high pathology), FN = False Negatives (dementia & low pathology).

**Supplementary Table 3. Clinical and demographic characteristics of Lewy body disease cohorts stratified by APOE status.**

| Cohort | n | Male, n (%) | Age at onset, years | Disease duration, years | Age at death, years | Dementia, n (%) | Orthostatic hypotension, n (%) | Ischaemic pathology, n (%) |
|---|---|---|---|---|---|---|---|---|
| APOE ε3 | 181 | 104/181 (57%) | 61.2 ± 11.9 | 16.6 ± 9.5 | 77.6 ± 8.1 | 97/181 (54%) | 58/84 (69%) | 56/181 (31%) |
| APOE ε4 | 68 | 48/68 (71%) | 60.5 ± 11.2 | 13.9 ± 7.9 | 74.4 ± 7.7 | 42/68 (62%) | 28/35 (80%) | 19/68 (28%) |
| **Cohort** | **Lewy body density (LB/mm²)** | **%Aβ** | **%pTau** | **α-synuclein Braak stage[3]** | **Aβ Thal phase[2]** | **pTau Braak & Braak stage[1]** | **ADNC[4, 5]** | **TDP-43 LATE stage[6]** |
| APOE ε3 | 1.749 [0.011–47.453] | 0.062 [0.001–7.846] | 0.508 [0.005–14.81] | 6 [0–6] | 1 [0–5] | 2 [0–6] | 1 [0–3] | 20/121 (17%) |
| APOE ε4 | 4.533 [0.046–34.998] | 1.351 [0.004–10.623] | 0.537 [0.007–18.803] | 6 [5–6] | 3 [0–5] | 2 [0–6] | 1 [0–3] | 7/44 (16%) |

Values are mean ± SD for age at onset, disease duration and age at death. Age at onset was calculated from reported symptom onset. Disease duration is symptom onset to death. Lewy body density, % area of Aβ (%Aβ), and % area of pTau (%pTau) are quantitative averages across defined cortical and medial temporal regions. Values for Lewy body density, %Aβ, and %pTau, α-synuclein Braak stage, Aβ Thal phase, pTau Braak & Braak stage and and Alzheimer's disease neuropathological change (ADNC) National Institute on Aging-Alzheimer's Association (NIA-AA)levels are median with [range]. ADNC: 0-3; Braak stage: 0-6; Aβ Thal: 0-5; pTau Braak & Braak stage: 0-VI (numbered 0-6). TDP-43 LATE staging represented as % cases, since all positive cases correspond to stage 2.

**Supplementary Table 4. Clinical and demographic characteristics of Lewy body disease cohorts stratified by dementia and APOE status.**

| Cohort | n | Male, n (%) | Age at onset, years | Disease duration, years | Age at death, years | Dementia, n (%) | Orthostatic hypotension, n (%) | Ischaemic pathology, n (%) |
|---|---|---|---|---|---|---|---|---|
| PD APOE ε3 | 84 | 44/84 (52%) | 61.3 ± 12.5 | 16.5 ± 9.5 | 77.3 ± 8.6 | 0/84 (0%) | 24/37 (65%) | 19/84 (23%) |
| PD APOE ε4 | 26 | 16/26 (62%) | 60.4 ± 10.9 | 14.1 ± 5.3 | 74.5 ± 7.9 | 0/26 (0%) | 10/13 (77%) | 9/26 (35%) |
| PDD/DLB APOE ε3 | 97 | 60/97 (62%) | 61.2 ± 11.3 | 16.7 ± 9.5 | 77.8 ± 7.6 | 97/97 (100%) | 34/47 (72%) | 37/97 (38%) |
| PDD/DLB APOE ε4 | 42 | 32/42 (76%) | 60.5 ± 11.5 | 13.8 ± 9.3 | 74.3 ± 7.6 | 42/42 (100%) | 18/22 (82%) | 10/42 (24%) |
| **Cohort** | **Lewy body density (LB/mm²)** | **%Aβ** | **%pTau** | **α-synuclein Braak stage[3]** | **Aβ Thal phase[2]** | **pTau Braak & Braak stage[1]** | **ADNC[4, 5]** | **TDP-43 LATE stage[6]** |
| PD APOE ε3 | 0.959 [0.011–12.627] | 0.041 [0.002–4.954] | 0.272 [0.005–7.23] | 6 [0–6] | 1 [0–4] | 2 [0–4] | 1 [0–2] | 6/51 (12%) |
| PD APOE ε4 | 0.87 [0.046–21.779] | 0.248 [0.004–4.143] | 0.266 [0.007–2.686] | 6 [5–6] | 3 [0–5] | 2 [0–4] | 1 [0–2] | 2/16 (13%) |
| PDD/DLB APOE ε3 | 3.352 [0.034–47.453] | 0.285 [0.001–7.846] | 0.811 [0.007–14.81] | 6 [0–6] | 2 [0–5] | 2 [0–6] | 1 [0–3] | 14/70 (20%) |

| | | | | | | | | |
|---|---|---|---|---|---|---|---|---|
| PDD/DLB APOE ε4 | 5.864 [1.046–34.998] | 2.397 [0.02–10.623] | 0.761 [0.039–18.803] | 6 [5–6] | 3 [0–5] | 2 [1–6] | 1 [0–3] | 5/28 (18%) |

Values are mean ± SD for age at onset, disease duration and age at death. Age at onset was calculated from reported symptom onset. Disease duration is symptom onset to death. Lewy body density, % area of Aβ (%Aβ), and % area of pTau (%pTau) are quantitative averages across defined cortical and medial temporal regions. Values for Lewy body density, %Aβ, and %pTau, α-synuclein Braak stage, Aβ Thal phase, pTau Braak & Braak stage and and Alzheimer's disease neuropathological change (ADNC) National Institute on Aging-Alzheimer's Association (NIA-AA)levels are median with [range]. ADNC: 0-3; Braak stage: 0-6; Aβ Thal: 0-5; pTau Braak & Braak stage: 0-VI (numbered 0-6). TDP-43 LATE staging represented as % cases, since all positive cases correspond to stage 2.

**Supplementary Table 5. Clinical and demographic characteristics of Lewy body disease cohorts stratified by TDP-43 LATE stage.**

| Cohort | n | Male, n (%) | Age at onset, y | Disease duration, y | Age at death, y | APOE ε4 carriers, n (%) | Dementia, n (%) | Orthostatic hypotension, n (%) |
|---|---|---|---|---|---|---|---|---|
| TDP-43 LATE stage 0 | 232 | 134/232 (58%) | 59.9 ± 11.5 | 17.0 ± 9.2 | 78.4 ± 8.0 | 41/176 (23%) | 100/232 (43%) | 84/121 (69%) |
| TDP-43 LATE stage 2 | 34 | 21/34 (62%) | 62.4 ± 12.6 | 18.1 ± 9.7 | 81.7 ± 7.9 | 7/30 (23%) | 21/34 (62%) | 7/20 (35%) |

| Cohort | Lewy body density (LB/mm$^2$) | %Aβ | %pTau | α-synuclein Braak stage[3] | Aβ Thal phase[2] | pTau Braak & Braak stage[1] | ADNC[4, 5] | Ischaemic pathology, n (%) |
|---|---|---|---|---|---|---|---|---|
| TDP-43 LATE stage 0 | 1.476 [0.002–34.998] | 0.28 [0.001–11.205] | 0.791 [0.005–18.803] | 6 [0–6] | 2 [0–5] | 2 [0–6] | 1 [0–3] | 0/232 (0%) |
| TDP-43 LATE stage 2 | 3.83 [0.003–47.453] | 0.892 [0.001–10.623] | 2.569 [0.011–19.313] | 6 [0–6] | 3 [0–5] | 2 [0–6] | 1 [0–3] | 34/34 (100%) |

Values are mean ± SD for age at onset, disease duration and age at death. Age at onset was calculated from reported symptom onset. Disease duration is symptom onset to death. Lewy body density, % area of Aβ (%Aβ), and % area of pTau (%pTau) are quantitative averages across defined cortical and medial temporal regions. Values for Lewy body density, %Aβ, and %pTau, α-synuclein Braak stage, Aβ Thal phase, pTau Braak & Braak stage and and Alzheimer's disease neuropathological change (ADNC) National Institute on Aging-Alzheimer's Association (NIA-AA) levels are median with [range]. ADNC: 0-3; Braak stage: 0-6; Aβ Thal: 0-5; pTau Braak & Braak stage: 0-VI (numbered 0-6). TDP-43 LATE staging represented as % cases, since all positive cases correspond to stage 2.

**Supplementary Table 6. Measurements collected for each pseudo-cell in Lewy body classifier.**

| Feature | Pseudo-cell compartment | Measurement |
|---|---|---|
| Intensity | Nucleus (original detection object), cytoplasm (2 µm background around detection object), cell (nucleus + cytoplasm) | Mean, median, min, max, and standard deviation for red, green, blue, deconvoluted haematoxylin, deconvoluted DAB, and deconvoluted residual channels. |
| Shape/morphology | Nucleus (original detection object) | Area, length, circularity, solidity, max diameter, min diameter. |
| Haralick[18] | Nucleus (original detection object) | Angular second moment (F0), contrast (F1), correlation (F2), sum of squares (F3), inverse difference moment (F4), sum average (F5), sum variance (F6), sum entropy (F7), entropy (F8), difference variance (F9), difference entropy (F10), information measure of correlation 1 (F11), information measure of correlation 2 (F12). |
